# Normative Modelling of Brain Volume in Multiple Sclerosis

**DOI:** 10.1101/2025.09.14.25335702

**Authors:** Max Korbmacher, Ingrid Anne Lie, Kristin Wesnes, Eric Westman, Thomas Espeseth, Ole Andreas Andreassen, Lars T. Westlye, Stig Wergeland, Hanne Flinstad Harbo, Gro Owren Nygaard, Kjell-Morten Myhr, Einar August Høgestøl, Øivind Torkildsen, the Alzheimer’s Disease Neuroimaging Initiative

## Abstract

**Background and Objectives:** Interpretation of brain atrophy in multiple sclerosis (MS) relies on group-level research and lacks individualized reference standards. Normative modelling can enable patient-level assessments of regional brain volumes relative to population expectations.

**Methods:** We constructed age-, sex-, and intracranial volume–adjusted normative models of regional cortical and subcortical FreeSurfer-estimated brain volumes and applied to data from a concluded clinical trial and routine hospital examinations to derive regional deviation Z-scores and counts of critical deviations (Z < –1.96). Associations with disability (Expanded Disability Status Scale [EDSS]), cognitive performance (Paced Auditory Serial Addition Test [PASAT]), and fatigue (Fatigue Severity Scale [FSS]) were examined cross-sectionally and longitudinally using fixed and random effects models. A deviation-based stratification rule was evaluated for disability progression and relapse risk using survival analyses.

**Results:** Models were trained on 62,444 MRI datasets from healthy individuals across the lifespan (50.8% females, age range 6.0-90.1) and applied to 953 longitudinal MRI scans from 362 people with MS (mean age = 38.8±9.7, 70.5% females, follow-up up to 12 years). People with MS exhibited a higher number of critical deviations than matched controls (incidence rate ratio 2.70, 95% CI 2.21–3.30), most prominently in the thalamus (approximately 25% of patients). A higher number of deviations was associated with higher disability (EDSS) indicated by a cross-sectional (β=0.24, 95% CI 0.14–0.34) and longitudinal main effect (β=0.07, 95% CI 0.02–0.13). Lower-than-reference volumes in the thalamus, hippocampus, and putamen were consistently associated with higher disability cross-sectionally (β_standardized_ = –0.17 to –0.23) and over time (β_standardized_ = –0.14 to –0.18). Deviation-based risk stratification identified patients with modestly higher disability trajectories (β=0.13, 95% CI 0.03–0.24).

**Discussion:** Normative modelling reveals a heterogeneous morphometric deviation profile in MS, centred on deep grey matter structures and associated with disability accumulation. These findings support the use of population-referenced MRI metrics for individual-level phenotyping in MS and warrant validation in independent cohorts.

## 1. Introduction

Interpreting biological measurements at the individual level remains a fundamental challenge across medicine and neuroscience, where population-level variability often obscures clinically meaningful deviations. Yet, imaging-based reference values are rarely applied in neurological practice, as technical and biological variability limit comparability across studies. Recent advances in normative modelling have begun to address these challenges, enabling the translation of population-level neuroimaging data into neuroscientific and clinical contexts.^1,2^ Normative models quantify the degree to which an individual measure deviates from the distribution of normative reference populations.^1^ Conventional volumetric approaches quantify absolute or group-level differences but do not indicate whether a given patient’s anatomy is unusually low or within expected normative ranges. Normative modelling directly addresses this gap by providing age-, sex-, and ICV-adjusted deviation scores that are interpretable at the individual level. Previous studies have established normative models first for whole brain volumes^3^ and then for common metrics of cortical thickness and surface area^1,4,5^ as well as subcortical volumes,^6^ yet comprehensive models that jointly characterise both cortical and subcortical regional volumes with direct clinical applicability are still limited.

Multiple sclerosis (MS) offers a well-characterised disease context in which to evaluate whether normative modelling can reveal concordant patterns of grey matter deviation. This is supported by previous studies presenting MS-specific cutoff-values for brain atrophy^7^ and highlighting the potential of reference values specific to MS to better explain disability.^8^ Over the past decades, magnetic resonance imaging (MRI) research in MS has evolved from a primary focus on lesion detection to the recognition of brain atrophy as a central marker of disease progression. Atrophy occurs across MS phenotypes and exceeds the rates observed in normal ageing,^9–11^ though technical limitations restricted its application for short-term individual monitoring.^12^ While deep grey matter, and particularly thalamic atrophy, presented associations with disability accumulation,^13^ cortical atrophy has been associated with cognitive decline.^10^ Alongside these advances, the broader clinical role of MRI has expanded beyond diagnosis to monitoring and treatment guidance, reflecting a shift in the field from viewing MRI primarily as a diagnostic tool to using it as a window into the long-term mechanisms of neurodegeneration and ageing in MS.^14^ However, these findings are largely based on group-level analyses and therefore provide limited guidance for interpreting individual patient scans, representing a major barrier to clinical translation. The next step is to establish generalisable, practically usable reference distributions against which individual scans can be contextualised.

Normative modelling directly addresses this clinical interpretability gap by estimating individual deviation profiles relative to age- and sex-adjusted reference distributions, thereby enabling a) rigorous testing of imaging markers proposed by the literature (e.g., cortical and thalamic atrophy),^9–11^ and b) discovery of additional, potentially overlooked markers that may not be apparent to standard neuroradiological assessment. By aggregating single-subject deviation profiles, one can quantify heterogeneity across people with MS (pwMS), delineate a reproducible morphometric phenotype characterised by coordinated deviations across distributed grey matter systems, and assess its clinical relevance for disability and progression.^15^ Such a framework is disease-agnostic and scalable: it aims to translate population-level neuroimaging into individualised brain phenotypes that are interpretable in clinical settings.

Here, we develop large-scale normative models for regional cortical and subcortical grey matter volumes derived from brain MRI and apply them to a pooled MS cohort to establish a morphometric phenotype of MS at the individual level. Using a reference dataset of N = 62,444 healthy controls (51% female) to derive age- and sex-adjusted expectations, we assess norm-deviations in N = 362 pwMS across 953 T1-weighted MRI scans spanning up to 12±3.72 years. We address three goals: i) to determine whether MS expresses a robust morphometric phenotype reflected by concordant deviations across deep and cortical grey matter; ii) to characterise heterogeneity in deviation profiles and quantify the extent of lower-than-reference regional volumes at the person level; and iii) to establish the clinical relevance of these deviations for disability and disease course, cross-sectionally and longitudinally. Together, this work provides a generalisable framework for individualised, reference-based interpretation of brain structure that is directly extensible to other neurological and psychiatric conditions.

## 2. Methods

### 2.1 Participants

#### 2.1.1 Healthy control cohort

We used seven international databases to assemble a large cross-sectional healthy control (HC) cohort (originally: N=63,115, N=62,795 after removal of missing values; for information see Supplemental Table 1), of which N=351 were used to one-on-one age-, sex-, and intracranial volume (ICV)-match a longitudinal MS dataset (N=351 with available MRI data; 82.8% females; age range 18.5-67.6 years; scans=953; median follow-up time=3.72 years, mean absolute deviation=3.75) using propensity scores at baseline. The remaining N=62,444 HC (50.8% females, age range 6.0-90.1) were used for model training.

#### 2.1.2 Multiple sclerosis cohort

The MS cohort was assembled from two independent datasets collected across Norway diagnosed using the McDonald criteria.^16^ The first MS sample included 88 pwMS who participated in the omega-3 fatty acid in MS (OFAMS) multicentre clinical trial conducted between 2004-2008^17^. The trial entailed monthly MRI acquisition, and clinical examination performed every 6 months over a 2-year period, followed by a single follow-up visit about 10 years after the original trial concluded (i.e. 25 brain scans per patient). The attrition rate was low with 96.6% (85 of 88) completing the 10-year follow-up. The second MS dataset was collected at the Oslo university hospital (OUH) during clinical and study assessments since 2012 with standard follow-ups (N=302, T_1_w-scans=690, mean time between first and final scan = 1.68±2.22 years), with recruitment at the first assessment.^18^ Both MS cohorts were clinically assessed for motor and cognitive impairments by evaluating a) disability, using the Expanded Disability Status Scale (EDSS),^19^ b) cognitive function, using Z-scores of the 3 second Paced Auditory Serial Addition Test (PASAT),^20^ and c) the level of fatigue, using the mean Fatigue Severity Scale (FSS) score.^21^ EDSS scores were available at all MRI timepoints, however, there was systematic missingness of PASAT and FSS scores in parts of the cohort, as these assessments were not used at all recordings. No other cognitive measures were available across time points and cohorts.

#### 2.1.1 Standard Protocol Approvals, Registrations, and Patient Consents

All data collections and usage were approved by respective ethical review boards, and informed consent forms were obtained (see Supplemental Note 1).

### 2.2 Magnetic resonance imaging (MRI)

#### 2.2.1 MRI acquisition

T_1_-weighted MRI data were obtained using various scanners, sites, and field strengths (1.5T or 3T). Acquisition protocols and machines were stable over time, however, due to the long-follow up time of the multi-centre study (OFAMS), scanner software was updated during the study period. An overview of the acquisition protocols and MRI sequence information can be retrieved from the original studies (HC: Supplemental Note 6, MS OFAMS data: Supplemental Table 2, original studies^17,18^).

MRI scans in the OFAMS cohort were acquired across multiple sites using scanners from different vendors and with heterogeneous protocols. As summarised in Supplemental Table 2, the study included both 1.5T and 3T systems from Siemens, Philips, and Toshiba, with considerable variation across sites in sequence parameters, including TR, TE, TI, flip angles, and voxel sizes. This applies to both the T1-weighted MPRAGE sequences and the FLAIR acquisitions. Some sites used 3D FLAIR sequences with approximately 1 mm voxel dimensions, while others used non-isotropic, thicker-slice implementations consistent with the original OFAMS trial report. Because OFAMS was a multicentre study conducted over several years, these differences reflect the site-specific protocols in place at the time. While MRI systems varied for OFAMS data, all data were collected on a single scanner in Oslo using a 3D sagittal brain volume (BRAVO) sequence for pre- and post-gadolinium contrast agent administration (1×1×1 mm resolution, TR = 8.16 ms, TE = 3.18 ms, TI = 450 ms, flip angle (FA) = 12°), and a 3D FLAIR sequence (1×1×1.2 mm resolution, TR = 8000 ms, TE = 127.25 ms, TI = 2240 ms) using a Discovery MR750 MRI (GE Medical Systems).

#### 2.2.2 MRI processing

T_2_-lesions were segmented using SPM’s lesion segmentation tool. The resulting lesion masks were visually quality controlled, and then coregistered to the T_1_-weighted images, using 7 degrees of freedom, correlation ratio as the cost function, and trilinear interpolation. Lesion counts and volumes were extracted from binarized lesion masks using fslmaths (https://fsl.fmrib.ox.ac.uk/fsl/fslwiki/Fslutils). Prior to lesion filling, we removed gadolinium-enhancing regions by applying a high-intensity filter: any voxel with an intensity above the 98th percentile was excluded, capturing both lesions and other enhancing areas. T_2_-hyperintense lesion masks were used to fill longitudinally co-registered T_1_-weighted images with the lesion_filling function implemented in FSL (version 5.0.10) to improve volume measurements by reducing intensity contrast within known lesion areas. T_1_-weighted lesions, defined as T_1_GD+ enhanced lesions, were counted by two experienced neuroradiologists for each dataset and not included in the lesion filling mask. After lesion-filling, regional brain volumes were extracted using the longitudinal pipeline of FreeSurfer for the longitudinal data (v7.1.1 OFAMS and v6.0.0 for Oslo data) including pwMS, and the cross-sectional FreeSurfer pipeline for cross-sectional HC data (multiple versions, see Supplemental Table 1), and then averaged across the brain regions specified in the Desikan-Killiany atlas.^22^ For training data harmonisation, we applied Combat, which was originally developed for batch effects in laboratory samples^23^ and recently extended to neuroimaging data.^24^ To not introduce incorrect group differences through harmonizing the test data,^25^ particular in the context of the small number of subjects per scanner site,^26^ we did not harmonise the test data. This approach preserves biological variance in the test data and avoids introducing artificial group differences when the number of subjects per site is small. As an additional quality control measure, we report results from harmonised test data in the Supplement (Supplemental Figures 3-6 and 12-13, Supplemental Note 4).

### 2.3 Statistical analysis

#### 2.3.1 Model training

We used multivariate fractional polynomial regression,^27^ trained on regional brain volumes of the HC cohort. We selected this framework because it provides a flexible but parsimonious framework for capturing non-linear lifespan trajectories, has been validated in large normative modelling efforts such as CentileBrain, and avoids the overfitting and instability typically associated with higher-order polynomials or spline models. Models were trained for each sex separately to model differences in both intercept and curvature that are not well captured by simple covariate or interaction terms, and trained for each brain region (i), where regional brain volumes were predicted from a linear effect of FreeSurfer reconstruction-based ICV, to account for confounding effects of head size, a fractional polynomial (p) of age (with maximum degree m=2), and an error term (u), with b_0-n_ indicating the regression coefficients. The non-linear age-term provides flexibly for non-linear lifespan trajectories without overfitting.

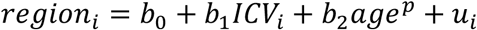

Model performance across regions was similar in training data R^2^ = 0.44±0.16, r = 0.66±0.13, root mean squared error (RMSE) (%) = 13.6±3.29 and mean absolute error (MAE) (%) = 10.6±2.58 compared to propensity matched HCs R^2^ = 0.34±0.19, r = 0.63±0.12, RMSE (%) = 14.80±3.96 and MAE (%) = 11.60±3.18. Note that for some small and/or partial-voluming affected regions with resulting poor/noisy signal,^28,29^ model performance was low, with the lowest observed R^2^=0.07 in the left temporal pole, followed by the right entorhinal cortex (R^2^=0.10). Results from these regions were interpreted with care and did not influence the main results or conclusions.

For an overview of region-wise model performance metrics see Supplemental Figure 7 and 8. Finally, to assess dataset-induced bias, we ran a leave-one-dataset-out analysis, where models were trained leaving out one of the utilised datasets (Supplemental Table 1) at a time. Across all regions, the Spearman correlations across all predictions were on average strong in training data (r = 0.99±0.02) and the propensity score matched HC (test) data (r = 0.99±0.03), used in case-control analyses, indicating robustness to dataset-specific effects.

#### 2.3.2 Associations between the number of deviations and clinical outcomes

The models were then used to predict the brain volumes per region in pwMS (N=362, T_1_w-scans=953), as well as in propensity-score matched cross-sectional HC sample (based on ICV, sex, and age, N=351). Z-scores, representing norm deviations, were calculated for each region from the true volumes, predicted individual-level volumes, and region-specific model error (RMSE).

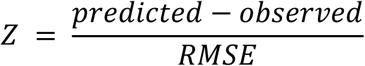

First, we quantified the total number of regional significant deviations or extreme lower-than-reference values, defined by Z < -1.96, defining critical deviations (CDs). A sensitivity analysis using a stricter threshold of Z < -2.33 is described in Supplemental Note 5. We then compared the number of CDs per individual between pwMS and HC across all regions (bilaterally), using negative binomial regression to account for overdispersion. The number of CDs across all regions (bilaterally) was then associated with EDSS, PASAT and FSS scores at baseline, using cross-sectional linear models, and longitudinally, using linear random intercept models, controlling for disease duration (DD):

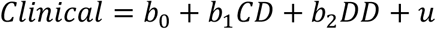

Note that, by training subgroup specific models and including ICV in model-training, the Z-scores in test data are already sex, age, and ICV-adjusted; however, we corrected for DD. Moreover, we did not include information on treatment in the modelling as the treatment plans were not comparable to each other and highly variable (Supplemental Tables 4-5). This was due to the historic developments of treatments and treatment strategies. While there were 119 relapses, this number was restricted to 43 participants during the entire study period.

Additionally, a large portion of these relapses were present during the first years of the OFAMS clinical trial (65 relapses across 37 patients) making this variable unsuitable as a covariate in longitudinal analyses. However, for quality control, we assessed the relationship between both Z-values and the number of CDs with the number of relapses at baseline, presenting no significant relationships (Supplemental Note 3).

There was a small increase of T_1_-weighted lesions over the first two years of the study period. A two-year period was chosen to allow for comparability between samples. There were only 2 new T_1_-weighted lesions in Oslo data and 18 in OFAMS data. The count of T_2_-weighted lesions fluctuated over time. We did not observe associations between T_2_-weighted lesions and Z-values or CDs, however significant associations were found for T_1_-weighted lesions (Supplemental Note 3). Note however that T_1_-weighted lesions were not a significant predictor of EDSS and did not improve model performance compared to the number of CDs (Supplemental Note 3). We did hence not include this variable in further analyses.

#### 2.3.3 Associations between the deviation magnitude (Z-scores) and clinical outcomes

Using the same logic from section 2.3.2, we assessed the impact of age, EDSS, PASAT and FSS on norm-deviations (Z) cross-sectionally and longitudinally using all regions (bilaterally).

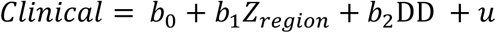

#### 2.3.4 Deviation profile–based stratification

To evaluate the clinical utility of normative deviation profiles, we implemented a deviation-based stratification approach. To re-iterate: A CD was defined as any regional brain volume with a Z-score < –1.96 relative to the normative reference distribution (section 2.3.2). To derive an interpretable stratification rule grounded in the empirical deviation patterns observed in Section 3.2, we identified the three brain regions with the highest overlap of CDs at baseline, referring to most commonly affected areas across pwMS, namely the bilateral thalamus, right superior parietal cortex, and pericalcarine cortex. We selected the three regions with the highest overlap of critical deviations because these exhibited a clear separation in overlap frequency: all three exceeded ≈14% participant-level overlap at baseline, while all other regions showed overlap values of ≈10% or lower. This provided an empirical cutoff for identifying the most consistently affected regions.

Participants were assigned to a deviation-based risk group if at least one CD was present in any of these regions at baseline. For bilateral structures, a deviation in either hemisphere qualified. Risk status was treated as a baseline characteristic and held constant for all longitudinal analyses.

Clinical relevance of deviation-based risk was assessed using a) linear mixed-effects models for EDSS, PASAT, and FSS with random intercepts for subjects and adjustment for disease duration (and age and sex where appropriate), and b) recurrent-event survival models for clinical relapses in the OFAMS sub-sample. Relapse dynamics were modelled using Andersen-Gill Cox models with robust variance estimates clustered by subject, with sensitivity analyses performed using Prentice-Williams-Peterson gap-time models. Effect estimates are reported as regression coefficients or hazard ratios with 95% confidence intervals, alongside model concordance (C-index).

#### 2.3.5 Reporting and power

For comparability, we report standardized regression coefficients or effect sizes. The significance-level was set at a conventional alpha level=0.05, and the Benjamini-Hochberg False Discovery Rate (FDR) correction for multiple comparisons^30^ was applied for all tests. MRI data were used when also data on sex and age were available. This allowed sex-specific predictions and calculation of Z-scores using the normative models. Missingness was addressed using multiple imputation, when missingness did not exceed 50% of the data or was non-random. We used multivariate imputation by chained equations, using a single iteration of predictive mean matching across 5 imputed datasets. Due to missingness, least data were available for cross-sectional correlations between FSS scores and brain volumes. Here, the minimal observable effect was f^2^=0.25 (corresponding to R^2^=0.20, and Pearson’s r=0.45), at 80% power, alpha=0.05, 3 nominator and 43 denominator degrees of freedom. For statistical analyses, R version 4.5.2 was used. To represent spatial statistics for the examined brain areas, we used the ggseg R package.^31^

## 3. Results

### 3.1 Descriptives

A total of 362 people with MS (pwMS) contributed 953 T1-weighted MRI sessions acquired longitudinally over the study period. At baseline, 351 pwMS with available MRI data matched to healthy controls were aged 38.6±9.7 years, of whom 250 (71.2%) were female, with a mean disease duration of 4.66±6.15 years, median EDSS of 2.0±0.7, PASAT score of 46.8±9.2, and mean FSS of 4.72±1.48. Across the full dataset, 72 EDSS scores (7.76%) were missing and were imputed, resulting in complete clinical data for all 953 MRI sessions. A total of 119 relapses were recorded across 43 participants during follow-up. Additional baseline characteristics for sub-samples are reported in Supplemental Table 3, with Z-score distributions and CDs shown in Supplemental Figures 1 and 11.

### 3.2 The morphometric profile of multiple sclerosis

In contrast to the population references, at baseline, the largest overlap of CDs across pwMS could be shown bilaterally in the thalami (25% and 26%, see Supplemental Figure 1 for distributions), followed by the right superior parietal cortex (16%) and pericalcarine (14%; Figure 1). Multiple other regions presented CDs in more than 10% of the pwMS, including the precuneus, fusiform area, putamen, inferior parietal area, posterior cingulate, hippocampus and parahippocampal area. These regions also presented the strongest deviations, measured by Z-scores (Supplemental Figure 1). In contrast, brain volumes among matched HC corresponded to reference levels (Supplemental Figure 2), and among the healthy population, the accumulation of CDs advanced slower (Supplemental Note 2). Examples for individual-level profiles for Z-scores can be found in Supplemental Figure 9 and for Z-score-based CDs in Supplemental Figure 10.

**Figure 1.**
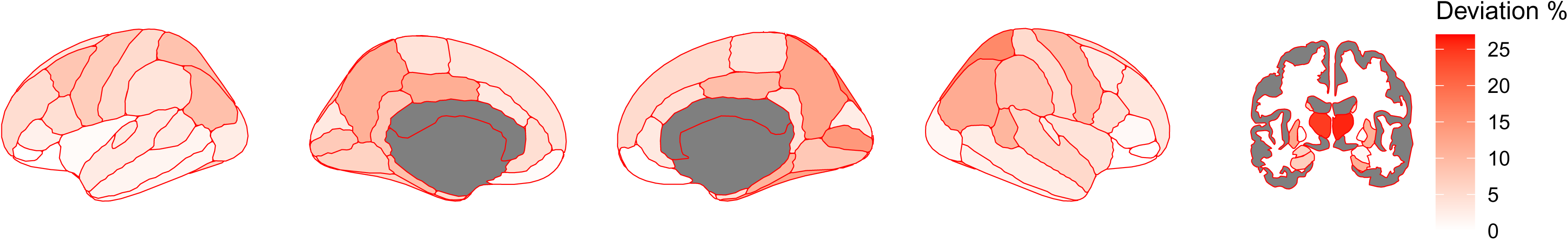
Overlap of significant deviations indicating lower brain volumes (Z<-1.96) across people with multiple sclerosis at baseline. Darker colours indicate a higher percentage of pwMS overlapping in lower-than-reference deviations in the respective brain region.

### 3.3 Associations between critical deviation count and clinical assessments

At baseline, additional CDs were associated with higher EDSS (β=0.24, 95% Confidence Interval (CI) [0.14; 0.34], p<0.00001), but not PASAT (p=0.155), FSS (p=0.131), or disease duration (p=0.727). Similarly, longitudinally, the number of CDs correlated with EDSS (β=0.07, 95% CI [0.02; 0.13], p = 0.016), but was not associated with PASAT (p=0.591), or FSS (p=0.255). Group-level cross-sectional case-control comparisons presented that pwMS had nearly three times the number of norm-deviations (4.51±4.95) compared to HC (1.67±2.67), indicated by an incidence rate ratio of 2.70, 95% CI [2.21; 3.30], p < 0.00001, and lower Z-scores than HC (average of regional mean absolute ΔZ=-0.22 (SE 0.48), d=-0.48, 95% CI [-0.17; -0.79], p=0.003). The regions with the largest negative Z-scores across pwMS, indicating smaller brain volumes, were the thalami (Z_left_=-1.16, Z_right_=-0.84), precuneus (Z_left_=-0.94, Z_right_=-0.94), posterior cingulate (Z_left_=-0.91, Z_right_=-0.70), and putamen (Z_left_=-0.87, Z_right_=-0.16).

### 3.4 Associations between the magnitude of norm-deviations and clinical assessments

At baseline, multiple regional deviations were significantly associated with EDSS and FSS, but not PASAT (p_FDR_>0.05; Figure 2). Significant associations were found between EDSS and volumetric deviations in cortical and subcortical regions, including the hippocampi (left: β_EDSS_=–0.17 95% CI [–0.27; –0.07]; p_FDR_=0.036; right: β_EDSS_=–0.21 95% CI [–0.31; –0.11], p_FDR_=0.004), right thalamus (right: β_EDSS_=–0.18 95% CI –0.29 to –0.07; p_FDR_=0.044), putamen (left: β_EDSS_=–0.21 95% CI [–0.31; –0.11], p_FDR_=0.004, right: β_EDSS_=–0.23 95% CI [– 0.33, –0.07], p_FDR_=0.002), and right amygdala (β_EDSS_=–0.17 95% [–0.28; –0.07], p_FDR_=0.036).

**Figure 2.**
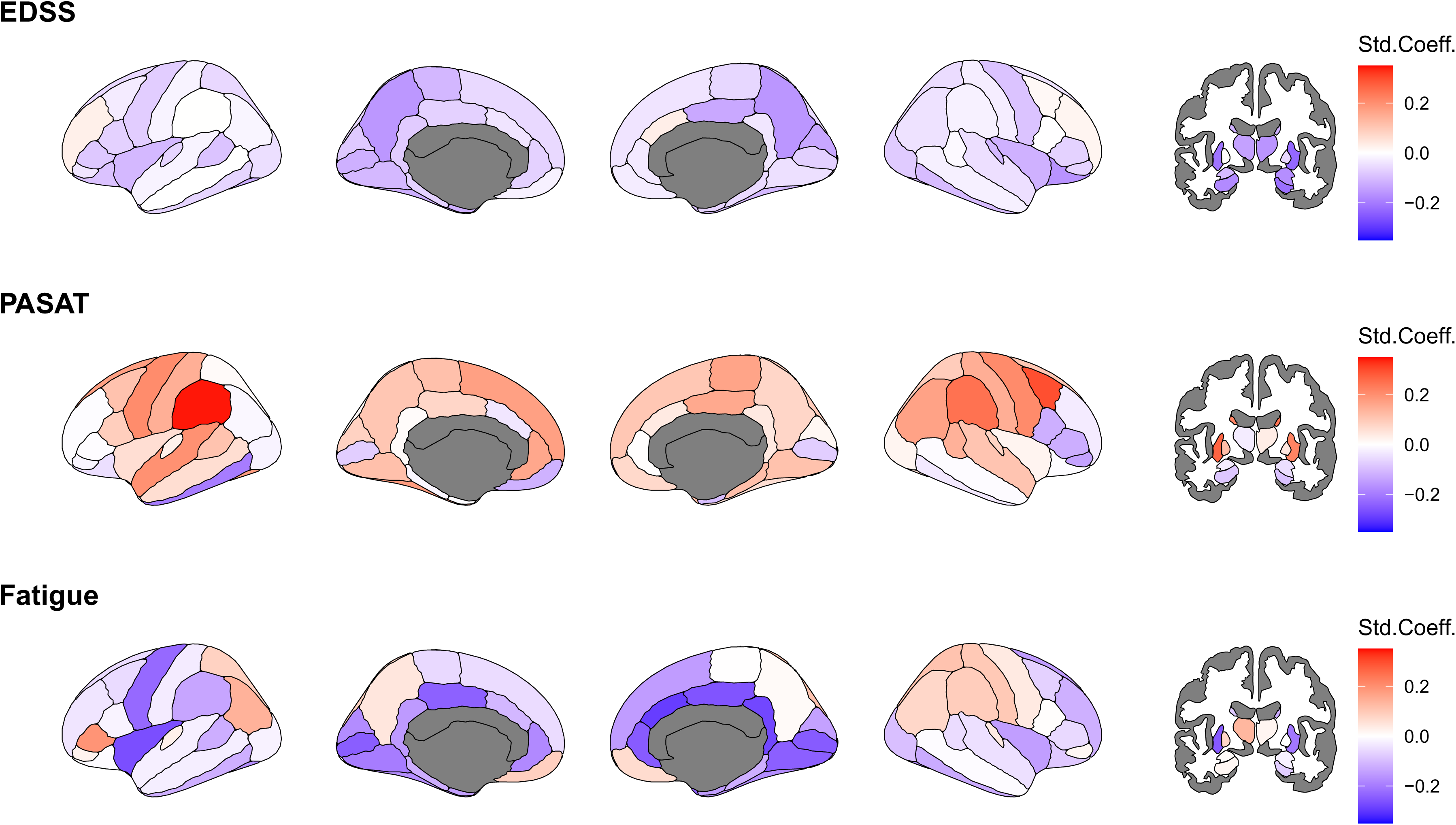
Baseline associations of age, EDSS, PASAT, and FSS on regional Z-scores. Std.Coeff.=standardized coefficient, EDSS= Expanded Disability Status Scale, PASAT= Paced Auditory Serial Addition Test, FSS=Fatigue Severity Scale. Note that only N=111 complete cases were available for PASAT and N = 119 for FSS scores. Considering the high level of missingness, imputation was not executed. Darker red colour indicates positive and darker blue colour negative norm-deviations, representing larger and smaller brain volumes compared to the reference. White colour indicates effects equal zero. Grey colour indicates uncorrected p>0.05.

Deviations from the cortical volumes in the left supramarginal gyrus (β_PASAT_=0.31 95% CI [0.13; 0.48], p_FDR_=0.010) predicted PASAT, and in the right isthmus cingulate (β_FSS_=–0.33 95% CI [–0.50; –0.16], p_FDR_=0.010) predicted FSS.

In longitudinal analyses, lower Z-values, indicating lower-than-reference brain volumes, affected widespread and yet function-specific areas, with significant relationships detected for EDSS (Figure 3). EDSS could be predicted by lower-than-reference volumes in the thalami (left: β_EDSS_=–0.15 95% CI [–0.23; –0.08]; p_FDR_=0.006; right: β_EDSS_=–0.14 95% [CI –0.22; –0.07]; p_FDR_=0.021), hippocampi (left: β_EDSS_=–0.16 95% CI [–0.22; –0.10]; p_FDR_<0.001; right: β_EDSS_=–0.15 95% CI [–0.22; –0.07]; p_FDR_=0.021), and the left putamen (β_EDSS_=–0.18 95% CI [–0.26; –0.11]; p_FDR_=0.043). No reference values could significantly predict FSS or PASAT scores.

**Figure 3.**
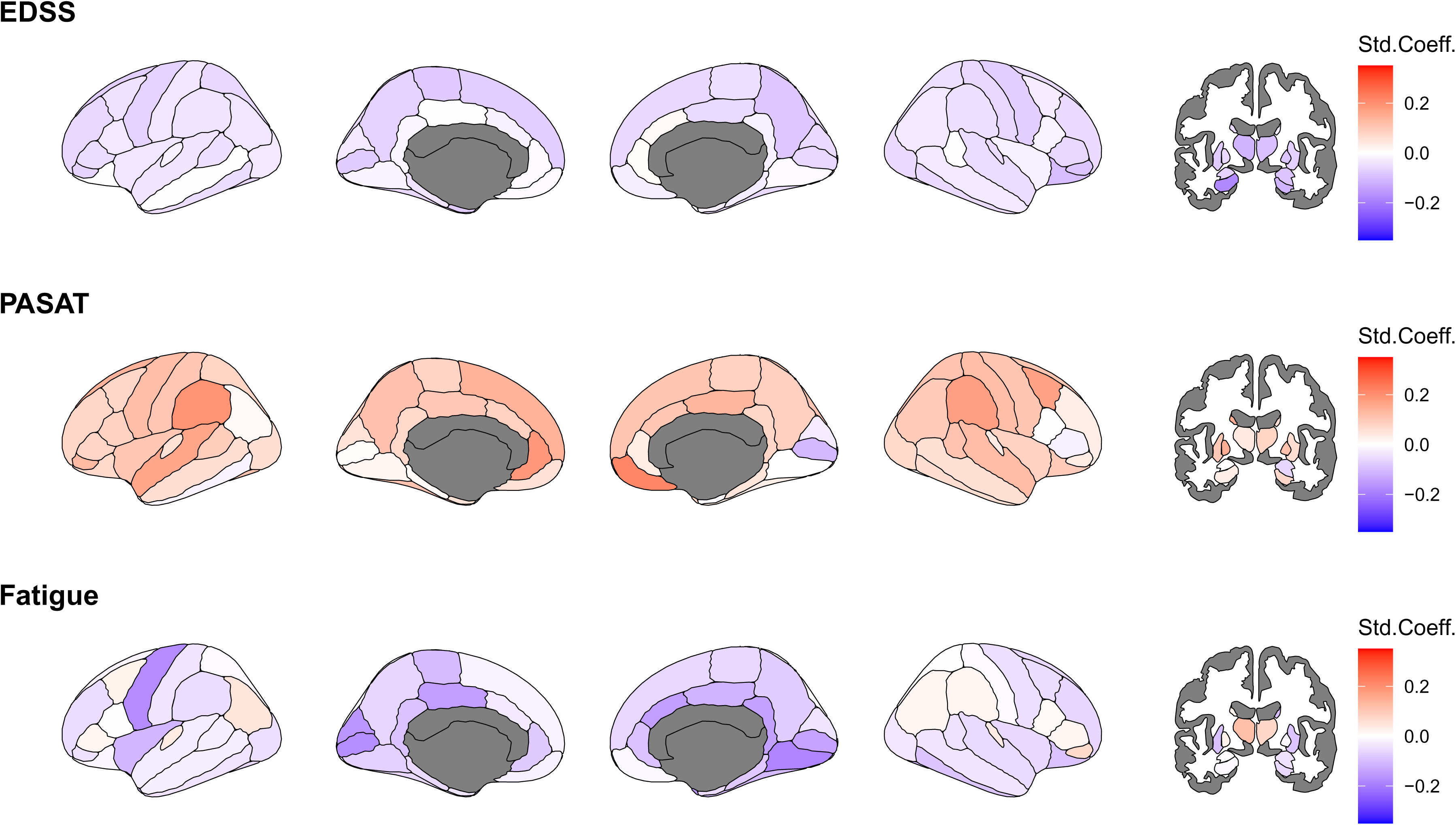
Longitudinal association of age, EDSS, PASAT, and FSS with regional Z-scores. Std.Coeff.=standardized coefficient, EDSS= Expanded Disability Status Scale, PASAT= Paced Auditory Serial Addition Test, FSS=Fatigue Severity Scale. Age and EDSS were available for all participants, for N=107 (222 sessions) for PASAT and N=110 (214) for FSS, with this high level of missingness not allowing for imputation. Darker red colour indicates positive and darker blue colour negative norm-deviations, representing larger and smaller brain volumes compared to the reference. White colour indicates effects equal zero. Grey colour present regions which were not assessed. All associations were significant *before* FDR-corrections. Corrected, significant relationships are reported in-text.

The results presented in sections 3.2-3.4 remained unchanged when applying a harmonisation strategy directly to the test data, except for counter-intuitive findings of lower levels of PASAT being associated with negative Z-scores (brain volumes smaller than the norm; see Supplemental Figures 3-6).

#### 3.4.5 Stratification based on morphometric profile

As the heterogeneous morphometric profile of MS was characterised by distinct regions, particularly the thalamus, superior parietal cortex and pericalcarine, CDs in these regions could be used as a stratification rule into a deviation-based risk group to indicate risk for disability, cognitive decline, fatigue and relapse accumulation.

##### 3.4.5.1 Group differences in EDSS, PASAT and FSS

Linear mixed models suggested higher EDSS over time in the deviation-based risk group (β_EDSS_ = 0.13 95% CI [0.03; 0.24], p = 0.01) when correcting for disease duration, but not PASAT (p = 0.43) or FSS (p = 0.41).

##### 3.4.5.2 Relapse risk and recurrence

To quantify the association between deviation-based risk group and relapse dynamics, we modelled time to relapse using survival analysis for recurrent events in the OFAMS sub-sample. In an Andersen-Gill Cox model with robust variance estimates clustered by subject, the presence of ≥1 CD was associated with an increased relapse hazard (hazard ratio [HR] = 1.60, 95% CI [0.93; 2.74]; Wald p = 0.09, Figure 4). Results were highly consistent in a Prentice-Williams-Peterson gap-time model stratified by event order, yielding a comparable effect estimate (HR = 1.61, 95% CI [0.94; 2.76]; robust p = 0.08). Model concordance was modest in both frameworks (AG: C = 0.54; PWP: C = 0.54), reflecting limited discrimination at the individual level. Together, these results indicate a stable, directionally consistent association between CDs and higher relapse risk across recurrent-event modelling approaches, although effect estimates did not reach conventional levels of statistical

**Figure 4.**
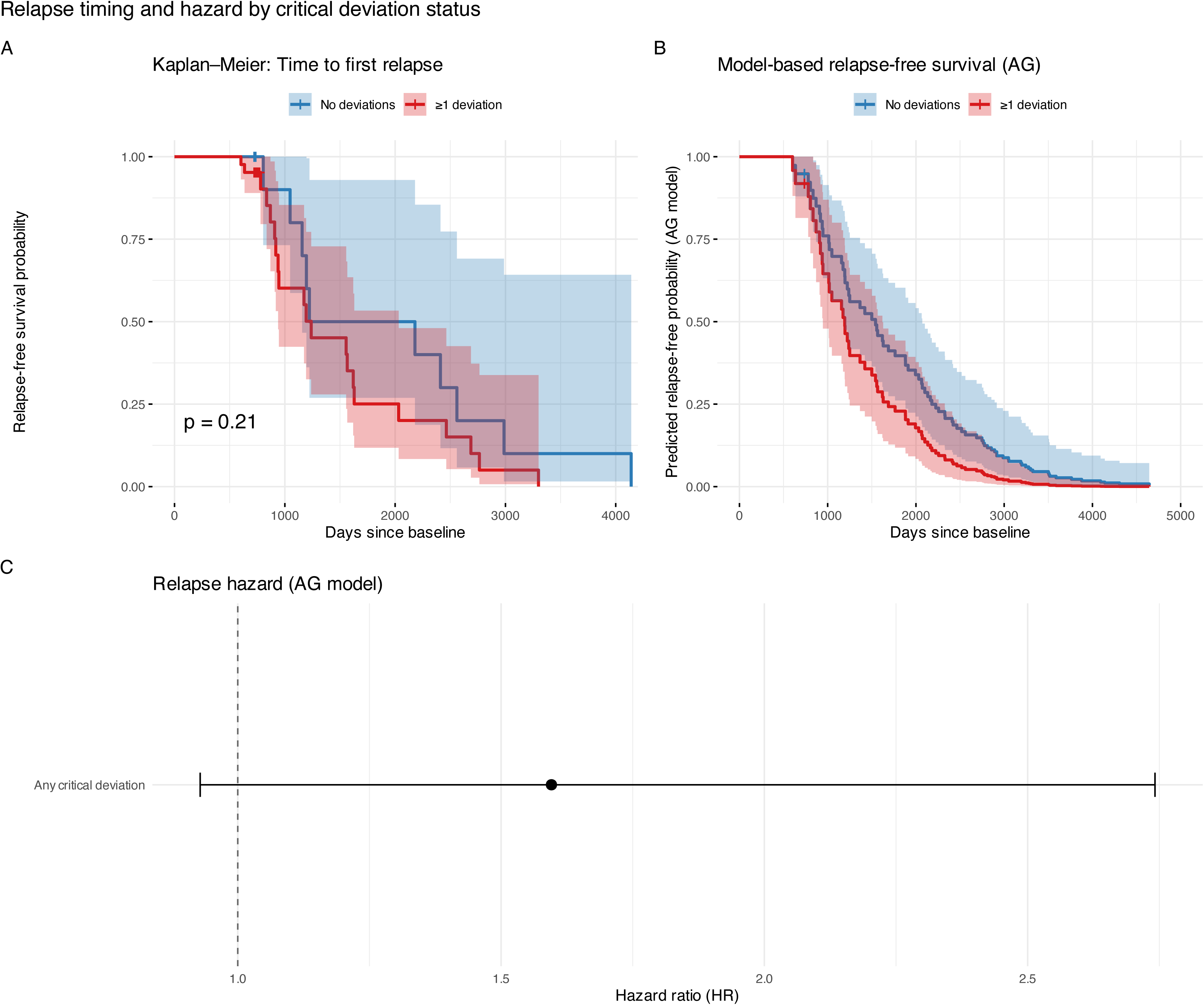
Relapse dynamics as a function of critical brain deviations. (A) Kaplan–Meier estimates of time to first clinical relapse, stratified by the presence of ≥1 critical deviation at baseline. Shaded areas indicate 95% confidence intervals. (B) Model-based relapse-free survival curves derived from an Andersen–Gill Cox model for recurrent events, comparing patients with and without critical deviations. (C) Forest plot summarising the hazard ratio for relapse associated with critical deviations from the Andersen–Gill model (points indicate hazard ratios; horizontal lines indicate 95% confidence intervals; dashed line denotes HR = 1). All Cox models used robust variance estimates clustered by subject to account for within-individual correlation.

## 4. Discussion

In this study, we applied large-scale normative models of regional brain volumes to longitudinal MRI data from pwMS and heterogeneous patterns of morphometric deviation relative to healthy population references. PwMS exhibited nearly three times the number of CDs compared with matched healthy controls. Among pwMS, CDs were unevenly distributed across regions, with the thalamus showing the largest and most consistent overlap across individuals. Approximately one quarter of the cohort exhibited critical thalamic deviations, highlighting this structure as the single most representative region within the MS morphometric profile. At the same time, deviation profiles varied markedly between individuals, underscoring the heterogeneity of grey matter involvement in MS. Both subcortical regions such as thalamus, putamen and limbic system, and occipital and parietal regions, such as superior parietal cortex, pericalcarine, precuneus, and fusiform area presented CDs in more than 10% of the pwMS. Both the magnitude of regional deviations, particularly in the thalamus, and the cumulative burden of CDs were associated with level of disability cross-sectionally and longitudinally. Lower-than-reference volumes in subcortical structures, particularly the thalamus, hippocampus, and putamen, were consistently linked to higher disability, and in frontal and cingulate cortical regions to lower cognitive performance and higher fatigue. Although associations between thalamic atrophy and disability are well established in MS,^9–11^ expressing these abnormalities as age- and sex-adjusted normative Z-scores provides interpretable, clinically implementable, individual-level metrics that extend beyond traditional group-level analyses.^1^ The morphometric profile also provided a basis for clinically interpretable stratification. By defining deviation-based risk using regions with the largest overlap of CDs, we identified patient subgroups with higher disability over time and a directionally consistent increase in relapse hazard. These findings illustrate how normative modelling can translate high-dimensional imaging data into compact risk indicators suitable for clinical contexts.

Longitudinal analyses indicated that divergence from normative expectations increased with time, suggesting that normative models may help disentangle disease-related neurodegeneration from normative ageing processes.^4,32^ A related point is that our findings do not contradict prior work reporting increasing contributions of ageing to atrophy in MS, but instead reflect a different modelling framework: by referencing each individual against large normative lifespan trajectories, we quantify deviations that persist even after accounting for expected age-related change. In this sense, our results complement earlier studies by showing that disease-related effects remain detectable beyond normative ageing and may follow distinct trajectories when evaluated relative to population expectations. Biologically, thalamic involvement may reflect degeneration of long-range axonal connections given its role as a hub for major white matter tracts, whereas frontal cortical deviations may reflect processes such as accelerated ageing or sustained neuroinflammation.^33^

Normative modelling offers several advantages over conventional volumetric analyses. By enabling individual-level inference, it can reveal subclinical abnormalities not apparent on visual inspection or group averaging,^34^ although formal clinical application will require further validation. Integrating deviation profiles with established MRI markers such as lesion burden and global atrophy measures may improve patient stratification and support future precision-medicine approaches in MS.^34,35^ The broader applicability of this framework also suggests potential utility in other neurological and neuropsychiatric disorders characterised by heterogeneous structural alterations.^36,37^

The application of normative models also opens avenues to better understand a) cases with diagnostic challenges, and b) the heterogeneity of disease trajectories in general. In diagnostically challenging cases, the individual-level Z-scores provide an objective reference to determine whether regional volumes are unusually low for a patient’s age and sex, supporting interpretation when MRI findings are subtle or ambiguous. Similar to previously reported deviation patterns in psychiatric disorders,^36,37^ we found extensive heterogeneity across pwMS, yet with larger region-specific overlaps than in psychiatric disorders (REFS?), especially in the thalamus. This underscores the potential of brain imaging assessments in MS and their further epidemiological investigation.

This study has several strengths, including the use of large, multi-site normative data, harmonised processing pipelines, robustness checks and independent longitudinal MS samples. As for limitations, normative models were trained on cross-sectional data, potentially limiting longitudinal inference. However, in contrast to other normative models such as BrainAge,^38–40^ the presented models could capture longitudinal processes related to disability progression. Additionally, brain-behaviour relationships are usually weak,^41^ which might be the reason for observed small and non-significant effects after multiple comparison correcting. Second, clinical data (particularly PASAT and FSS) were sparsely available and missing at random, limiting the statistical power for some analyses, precluding imputation, and reducing our ability to include additional clinically meaningful covariates in the analyses. Accordingly, certain additionally clinically meaningful analyses, such as progression independent of relapse activity grouping was not possible. Moreover, treatments evolved over time and hence, disease-modifying treatment (DMT) histories differed widely in timing, duration, switching patterns, and documentation quality, any coarse categorisation would introduce misclassification and bias; therefore, DMT effects were not modelled in the present analysis. Future studies with complete longitudinal assessments of cognition and fatigue and more diverse samples (beyond Norway) are needed to validate and extend these findings. Additionally, while we used validated segmentation pipelines, volume estimates may be affected by scanner and MRI protocol variability, particularly in longitudinal settings, as well as processing pipelines. Here, we used two different FreeSurfer versions (v7.1.1 and v6.0.0). Although discrepancies between FreeSurfer versions can affect volumetric estimates, the magnitude of these differences is considerably reduced for more recent releases, particularly those published in close temporal succession.^42^ Recent evidence also shows that normative models are robust to FreeSurfer version differences^1^ and version differences normative modelling approaches have been shown to be relatively robust to such variation.^43^ Nevertheless, fundamental limitations of the FreeSurfer pipeline remain, such as potential errors in volume estimates resulting from misalignments of regional misalignments with the selected atlas within and between individuals. When using the models proposed here for unseen data from various scanners, harmonisation using tools such as ComBat^23,24^ or HACA3^44^ are recommended.

Since we found an association between T_1_-weighted lesions and both Z-values and the number of CDs, we cannot differentiate whether the presence of lesions biases volumetric estimates in themselves, in turn, influencing the magnitude of the presented Z-scores, or whether there are biologically meaningful associations. However, CDs were stronger and significantly associated with EDSS, which was not the case for T_1_-weighted lesions, there was no significant interaction effect between EDSS and T_1_-weighted lesions, while multicollinearity was low (Supplemental Note 3). Still, our sensitivity analyses using harmonised data produced concordant trends, reinforcing the robustness of the primary findings (see Supplemental Note 4 for deviations). Finally, although the present approach demonstrates the potential value of normative modelling for understanding individual differences in MS, it is not yet suitable for clinical use. This study represents an initial methodological step, and further work is needed to establish generalisability across scanners and centres, including calibration procedures, image-level or statistical harmonisation, and the development of more advanced modelling frameworks such as Bayesian hierarchical approaches or MS-specific reference curves.^8^ Any future clinical translation will also require integration with regulatory-approved volumetric tools and validation in independent, prospectively collected multi-centre cohorts. Our intent here is therefore methodological rather than clinical, and we view this work as a foundation for more refined and validated applications in future datasets.

In summary, by embedding regional brain volumes within a normative reference framework, we demonstrate that MS is characterised by a robust and recurrent spatial pattern of affected regions at the individual level, yet heterogeneous at the group level, with this pattern being clinically meaningful at both levels. Concordant deviations, most prominently involving subcortical regions such as thalamus, putamen and limbic system, and occipital and parietal regions, relate to disability, cognitive function, and fatigue, and can be translated into compact deviation profiles that support patient stratification. While further validation in independent cohorts is required, these findings suggest that normative modelling provides a scalable and interpretable bridge between population-level neuroimaging and individual-level clinical assessment, with potential applications for early detection, monitoring, and personalised management of neurodegeneration in MS.

## Open Data and Materials

Summary statistics and utilized code can be found at https://osf.io/d6nfq. Trained models can be found at https://doi.org/10.17605/OSF.IO/6R8DY. Multiple of the utilized dataset are sensitive, require IRB approval for usage, and can therefore not be openly shared.

## Acknowledgments

We extend our sincere gratitude to the pwMS who participated in this study, whose contributions are invaluable. This research originated from different research efforts: one national intervention study (OFAMS) and ten years follow-up examinations, as well the ongoing data collections at the MS research group at the Oslo University Hospital.

We thank the OFAMS study group at numerous hospitals across Norway, as well as the group of clinicians and researchers at the Oslo University Hospital.

Finally, we want to extend our gratitude to the thousands of scanned participants in the UK Biobank study and the other utilized cohorts as well as facilitators who made this study possible by providing invaluable data to the research community.

## Contributions (CRediT)

MK: Conceptualisation, Data curation, Formal analysis, Investigation, Methodology, Project administration, Software, Visualization, Writing – original draft

IAL: Data curation, Writing – review & editing KW: Writing – review & editing

EW: Writing – review & editing TW: Writing – review & editing

OAA: Funding Acquisition, Resources, Writing – review & editing

LTW: Funding Acquisition, Resources, Writing – review & editing

SW: Writing – review & editing

HFH: Funding Acquisition, Resources, Writing – review & editing

GON: Writing – review & editing

KMM: Resources, Writing – review & editing

EAH: Funding Acquisition, Supervision, Writing – review & editing

ØT: Funding Acquisition, Resources, Supervision, Writing – review & editing

