## Supplement for "Normative Modelling of Brain Volume in Multiple Sclerosis"

Article: Normative Models of Brain Volume for Diagnostic and Prognostic Stratification in Multiple Sclerosis, Korbmacher et al. (2025).

### Contents

*Supplemental Tables* ___ *2*

Supplemental Table 1: Overview of the cross-sectional healthy control samples training data set ___ 2

Supplemental Table 2: Overview of the OFAMS (multiple sclerosis) sample scanners and acquisition protocols ___ 3

Supplemental Table 3: Baseline demographic and clinical characteristics of the pwMS cohorts. ___ 4

Supplemental Table 4: Overview of administered treatments in Oslo data. ___ 5

Supplemental Table 5: Overview of administered treatments in OFAMS (Bergen) data. ___ 7

*Supplemental Figures* ___ *10*

Supplemental Figure 1. Distributions of the number of extreme deviations and Z-scores of norm deviations in the thalami in pwMS (non-harmonised test data) ___ 10

Supplemental Figure 2. Region-averaged Z-values in MS and HC ___ 11

Supplemental Figure 3. Overlap of significant deviations indicating lower brain volumes (Z<-1.96) across participants at baseline for harmonised data ___ 12

Supplemental Figure 4. Region-averaged Z-values in MS and HC for harmonised data ___ 13

Supplemental Figure 5: Overlap of significant deviations indicating lower brain volumes (Z<-1.96) across participants at baseline for harmonised data ___ 14

Supplemental Figure 6. Longitudinal association of age, disability (EDSS), cognitive performance (PASAT), and fatigue on regional Z-scores for harmonised data ___ 15

Supplemental Figure 7. Model performance: Variance explained and correlation coefficients for training (internal) and propensity matched (external) data. ___ 16

Supplemental Figure 8. Model performance: Root mean squared error and mean absolute error for training (internal) and propensity matched (external) data. ___ 17

Supplemental Figure 9. Individual-level Z-scores for minimum and maximum EDSS and disease duration 18

Supplemental Figure 10. Individual-level Z-score-based extreme deviations for minimum and maximum observed EDSS and disease duration ___ 19

Supplemental Figure 11. Distribution of Z-scores at baseline in people with MS ___ 20

Supplemental Figure 12. Individual-level Z-scores for minimum and maximum EDSS and disease duration using harmonised test data ___ 21

Supplemental Figure 13. Individual-level Z-score-based extreme deviations for minimum and maximum observed EDSS and disease duration using harmonised test data ___ 22

*Supplemental Notes* ___ *23*

Supplemental Note 1. Ethics Approvals ___ 23

Supplemental Note 2. Longitudinal changes in training data with available longitudinal sessions. ___ 24

Supplemental Note 3. Quality control analyses: associations between a) relapses, lesions, and b) Z-scores and extreme deviations. ___ 25

Supplemental Note 4. Differences between main and sensitivity analyses. ___ 27

*References* ___ *31*

### Supplemental Tables

***Supplemental Table 1:*** **Overview of the cross-sectional healthy control samples training data set**

| **Data** | **Sample size** | **Mean age ± SD, range** | **FreeSurfer[1] version** |
|---|---|---|---|
| ABCD[2] | 11 279 | 11.9±0.6, 10.6-13.6 | 5.3.0 |
| AddNeuroMed[3] | 292 | 74.2±6.2, 53.0-90.1 | 5.3.0 |
| ADNI[4] | 60 | 71.1±6.2, 56.6-86.4 | 7.4.1 |
| Human Connectome Project[5] (HCP)[6] | 1 097 | 28.8±3.7, 22.0-37.0 | 5.3.0 |
| Rockland Sample[7] | 827 | 35.3±20.9, 6.0-85.0 | 7.1.0 |
| Thematically Organized Psychosis Research study [local Oslo data] | 972 | 33.1±10.2, 13.0-72.0 | 7.4.1 |
| UK Biobank[8] | 48 040 | 64.9±7.8, 44.6-83.7 | 5.3.0 |

Healthy control training sample after removing missing data and not including the propensity-matched datasets: n=62 414. Superscript references can be found in the reference list at the end of the document.

***Supplemental Table 2:*** **Overview of the OFAMS (multiple sclerosis) sample scanners and acquisition protocols**

| | **1 (3)** | **2 (13)** | **3 (3)** | **4 (1)** | **5 (3)** | **6 (4)** | **7 (3)** | **8 (5)** |
|---|---|---|---|---|---|---|---|---|
| Scanner | Siemens Aera | Siemens Prisma | Siemens Skyra | Siemens Avanto | Siemens Skyra | Siemens Avanto | Siemens Aera | Philips Achieva |
| Field strength | 1.5T | 3T | 3T | 1.5T | 3T | 1.5T | 1.5T | 1.5T |
| Sequences | T1; MPRAGE T2; FLAIR | T1; MPRAGE T2; FLAIR | T1; MPRAGE T2; FLAIR | T1; MPRAGE T2; FLAIR | T1; MPRAGE T2; FLAIR | T1; MPRAGE T2; FLAIR | T1; MPRAGE T2; FLAIR | T1; FFE T2; FLAIR |
| TR (ms) | 1940 5000 | 1800 5000 | 2300 5000 | 2060 5000 | 2300 5000 | 2200 6000 | 2200 5000 | 7.6 4800 |
| TE (ms) | 2.69 335 | 2.28 386 | 2.32 387 | 3.10 340 | 2.32 387 | 2.82 358 | 2.67 335 | 3.75 338 |
| TI (ms) | 976 1800 | 900 1800 | 900 1800 | 1100 1800 | 900 1800 | 900 2200 | 900 1800 | 1660 1650 |
| Flip angle (¬∞) | 8 120 | 8 120 | 8 120 | 15 120 | 8 120 | 8 120 | 8 120 | 8 90 |
| Voxel size (mm) | 1.00x0.98x 0.98 1.00x1.00x 1.00 | 1.00x1.00x 1.00 | 1.00x1.00x 1.00 | 1.00x1.00x 1.00 | 0.9x0.94x0. 94 0.9x0.45x0. 45 | 1.00x0.49x 0.49 1.00x0.51x 0.51 | 0.90x0.94x 0.94 0.90x0.45x 0.45 | 1.00x0.98x0.98 1.00x1.00x1.00 |
| **Protocol (N of patients)** | | **9 (12)** | **10 (2)** | **11 (6)** | **12 (1)** | **13 (2)** | **14 (8)** | **15 (3)** |
| Scanner | | Philips Achieva | Siemens Prisma | Philips Ingenia | Toshiba MRT200SP 3 | Philips Ingenia | Siemens Prisma | Philips Achieva |
| Field strength | | 3T | 3T | 1.5T | 1.5T | 3T | 3T | 1.5T |
| Sequences | | T1; FFE T2; FLAIR | T1; MPRAGE T2; FLAIR | T1; FFE T2; FLAIR | T1; FFE T2; FLAIR | T1; FFE T2; FLAIR | T1; MPRAGE T2; FLAIR | T1; FFE T2; FLAIR |
| TR (ms) | | 8.04 5000 | 1800 5000 | 25 4800 | 13.5 1160 | 11.11 4800 | 1800 5000 | 7.1 4800 |
| TE (ms) | | 3.68 386 | 2.28 385 | 9.21 367 | 5.50 105 | 6.29 296 | 2.26 387 | 2.2 307 |
| TI (ms) | | 900 1800 | 900 1800 | 1660 2300 | 1650 906 | 1800 | 1800 | 1660 |
| Flip angle (¬∞) | | 8 90 | 8 120 | 30 90 | 20 90 | 8 90 | 8 120 | 8 90 |
| Voxel size (mm) | | 1.00x0.98x 0.98 1.00x0.98x 0.98 | 1.00x0.46x 0.46 0.50x0.73x 0.73 | 1.00x0.50x 0.50 1.00x0.50x 0.50 | 1.00x0.94x 0.94 0.50x0.74x 0.74 | 0.39x0.65x 0.39 0.47x4.0x0. 47 | 0.90x0.67x 0.67 0.56x0.98x 0.98 | 1.00x1.000x1.00 1.00x1.000x1.00 1.00x1.00x1.00 0.76x1.00x1.00 |

***Supplemental Table 3:*** **Baseline demographic and clinical characteristics of the pwMS cohorts.**

| | Total Sample (N=362) | OFAMS (N=84) | OUH (N=302) |
|---|---|---|---|
| Age | 38.8 (9.7) | 39.8 (8.4) | 38.6 (9.9) |
| Female | 256 (70.5%) | 54 (64.3%) | 220 (72.8%) |
| Male | 107 (29.5%) | 30 (35.7%) | 82 (27.2%) |
| EDSS | 2.0 (0.7) | 2.0 (0.8) | 2.0 (0.7) |

Mean scores ± standard deviations are displayed for age and median scores ± mean absolute deviation for EDSS. Note that there was missingness at random for EDSS scores, and hence imputation was applied. The availability of other clinical assessments (PASAT and FSS) was limited at baseline, not allowing for imputation. MRI data were available for N=362 participants.

*Supplemental Table 4:* **Overview of administered treatments in Oslo data.**

| **Patient** | **Session 1** | **Session 2** |
|---|---|---|
| | Unknown | Unknown |
| 1 | Teriflunomide | Alentuzumab |
| 2 | Fingolimod | Alentuzumab |
| 3 | None | None |
| 4 | None | Glatiramer Acetate |
| 5 | Fingolimod | Alentuzumab |
| 6 | Fumarate | Dimethylfumarate |
| 7 | Fingolimod | Fingolimod |
| 8 | Alentuzumab | Alentuzumab |
| 9 | Alentuzumab | Alentuzumab |
| 10 | Fingolimod | Fingolimod |
| 11 | Glatiramer Acetate | Glatiramer Acetate |
| 12 | None | None |
| 13 | Glatiramer Acetate | Glatiramer Acetate |
| 14 | None | None |
| 15 | Glatiramer Acetate | NA |
| 16 | Teriflunomide | Teriflunomide |
| 17 | Glatiramer Acetate | NA |
| 18 | Teriflunomide | Glatiramer Acetate |
| 19 | Fingolimod | Fingolimod |
| 20 | Teriflunomide | Teriflunomide |
| 21 | Fingolimod | Teriflunomide |
| 22 | Fingolimod | Fingolimod |
| 23 | None | None |
| 24 | Teriflunomide | Teriflunomide |
| 25 | None | Teriflunomide |
| 26 | None | None |
| 27 | None | None |
| 28 | Glatiramer Acetate | Glatiramer Acetate |
| 29 | None | None |
| 30 | Glatiramer Acetate | NA |
| 31 | Fingolimod | Fingolimod |
| 32 | None | None |
| 33 | Fingolimod | Fingolimod |
| 34 | Natalizumab | Natalizumab |
| 35 | Alemtuzumab | Alemtuzumab |
| 36 | None | Teriflunomide |

| | | |
|---|---|---|
| 37 | Glatiramer Acetate | None |
| 38 | Glatiramer Acetate | Glatiramer Acetate |
| 39 | None | None |
| 40 | Teriflunomide | Teriflunomide |
| 41 | Glatiramer Acetate | NA |
| 42 | Teriflunomide | Teriflunomide |
| 43 | Interferon | Fingolimod |
| 44 | None | Teriflunomide |
| 45 | Fingolimod | Fingolimod |
| 46 | Alemtuzumab | Alemtuzumab |
| 47 | None | None |
| 48 | Teriflunomide | DimethylDimetylfumarate |
| 49 | None | NA |
| 50 | Fingolimod | Fingolimod |
| 51 | Dimetylfumarate | DimethylDimetylfumarate |
| 52 | Fingolimod | Fingolimod |
| 53 | Teriflunomide | Teriflunomide |
| 54 | Glatiramer Acetate | NA |
| 55 | Fingolimod | None |
| 56 | None | None |
| 57 | Teriflunomide | NA |
| 58 | None | NA |
| 59 | None | None |

*Supplemental Table 5:* **Overview of administered treatments in OFAMS (Bergen) data.**

| Patient | DMT1 Name | DMT2 Name | DMT3 Name | DMT4 Name | DMT5 Name |
|---|---|---|---|---|---|
| 1 | Rebif 44 mcg | Copaxone | Extavia | | |
| 2 | Rebif 44 mcg | | | | |
| 3 | Rebif 44 mcg | | | | |
| 4 | Rebif 44 mcg | | | | |
| 5 | Rebif 44 mcg | Copaxone | | | |
| 6 | Rebif 44 mcg | Copaxone | | | |
| 7 | Rebif 44 mcg | Rebif 44 mcg | Copaxone | | |
| 8 | | | | | |
| 9 | Rebif 44 mcg | | | | |
| 10 | Rebif 44 mcg | Copaxone | | | |
| 11 | Rebif 44 mcg | Tecfidera | | | |
| 12 | Rebif 44 mcg | Avonex | Copaxone | | |
| 13 | Rebif 44 mcg | | | | |
| 14 | Rebif 44 mcg | | | | |
| 15 | Rebif 44 mcg | Rebif 22 mcg | Copaxone | Gilenya | |
| 16 | Rebif 44 mcg | Tysabri | Gilenya | Tysabri | |
| 17 | Tecfidera | | | | |
| 18 | Rebif 44 mcg | Copaxone | Tysabri | HSCT Mexico | |
| 19 | Rebif 44 mcg | Copaxone | Tecfidera | | |
| 20 | Rebif 44 mcg | Tysabri | | | |
| 21 | Rebif 44 mcg | | | | |
| 22 | Rebif 44 mcg | | | | |
| 23 | Copaxone | | | | |
| 24 | Rebif 44 mcg | | | | |
| 25 | Rebif 44 mcg | Gilenya | | | |
| 26 | Rebif 44 mcg | Copaxone 20 mg | Tysabri | | |
| 27 | Rebif 44 mcg | | | | |
| 28 | Rebif 44 mcg | | | | |
| 29 | | | | | |

| | | | | | |
|---|---|---|---|---|---|
| 30 | Rebif 44 mcg | | | | |
| 31 | Rebif 44 mcg | | | | |
| 32 | | | | | |
| 33 | Avonex | Copaxone | Rebif 44 mcg | Tysabri | |
| 34 | Rebif 44 mcg | Rebif 22 mcg | | | |
| 35 | | | | | |
| 36 | | | | | |
| 37 | Rebif 44 mcg | | | | |
| 38 | Rebif 44 mcg | Rebif 44 mcg | Aubagio | | |
| 39 | Rebif 44 mcg | Copaxone | Tysabri | | |
| 40 | Rebif 44 mcg | Aubagio | | | |
| 41 | Rebif 44 mcg | Aubagio | | | |
| 42 | Rebif 44 mcg | Rebif 44 mcg | Aubagio | | |
| 43 | Rebif 44 mcg | Copaxone | Tysabri | Tecfidera | Gilenya |
| 44 | | | | | |
| 45 | Rebif 44 mcg | | | | |
| 46 | Rebif 44 mcg | Copaxone | | | |
| 47 | Rebif 44 mcg | Copaxone | | | |
| 48 | Rebif 44 mcg | Avonex | | | |
| 49 | Rebif 44 mcg | | | | |
| 50 | Rebif 44 mcg | Tecfidera | | | |
| 51 | Rebif 44 mcg | Copaxone | Gilenya | | |
| 52 | | | | | |
| 53 | Rebif 44 mcg | | | | |
| 54 | Rebif 44 mcg | Avonex | Tysabri | Gilenya | |
| 55 | Rebif 44 mcg | | | | |
| 56 | Rebif 44 mcg | Copaxone 20,g | Tysabri | HSCT Moskva | |
| 57 | Rebif 44 mcg | Copaxone | | | |
| 58 | Rebif 44 mcg | Betaferon | | | |
| 59 | Rebif 44 mcg | Rebif 22 mcg | | | |
| 60 | Rebif 44 mcg | | | | |
| 61 | Rebif 44 mcg | Mitoxantrone | | | |

| | | | | | |
|---|---|---|---|---|---|
| 62 | Rebif 44 mcg | | | | |
| 63 | Rebif 44 mcg | Avonex | Tecfidera | | |
| 64 | Rebif 44 mcg | Copaxone | Avonex | | |
| 65 | Rebif 44 mcg | Tysabri | | | |
| 66 | Rebif 44 mcg | Tecfidera | | | |
| 67 | Rebif 44 mcg | Rebif 22 mcg | Copaxone | Tecfidera | |
| 68 | Rebif 44 mcg | | | | |
| 69 | Rebif 44 mcg | Copaxone | | | |
| 70 | Rebif 44 mcg | Copaxone | | | |
| 71 | Rebif 44 mcg | Rebif 22 mcg | Copaxone | | |
| 72 | Rebif 44 mcg | Copaxone 20 ,g | Aubagio | | |
| 73 | Rebif 44 mcg | Rebif 22 mcg | | | |
| 74 | Rebif 44 mcg | IVIG | Gilenya | Tysabri | IVIG |
| 75 | Rebif 44 mcg | Tysabri | | | |
| 76 | Avonex | Tysabri | Gilenya | | |
| 77 | Rebif 44 mcg | Tecfidera | Gilenya | | |
| 78 | Rebif 44 mcg | Rebif 22 mcg | Copaxone | | |
| 79 | Rebif 44 mcg | Rebif 22 mcg | Copaxone | | |
| 80 | Rebif 44 mcg | Copaxone 20 mg | | | |
| 81 | Rebif 44 mcg | Copaxone | | | |
| 82 | | | | | |
| 83 | Rebif 44 mcg | | | | |
| 84 | Rebif 44 mcg | Copaxone 20 mg | Copaxone 40 mg | Aubagio | |
| 85 | Rebif 44 mcg | Copaxone | | | |

### Supplemental Figures

***Supplemental Figure 1.*** **Distributions of the number of extreme deviations and Z-scores of norm deviations in the thalami in pwMS (non-harmonised test data)**

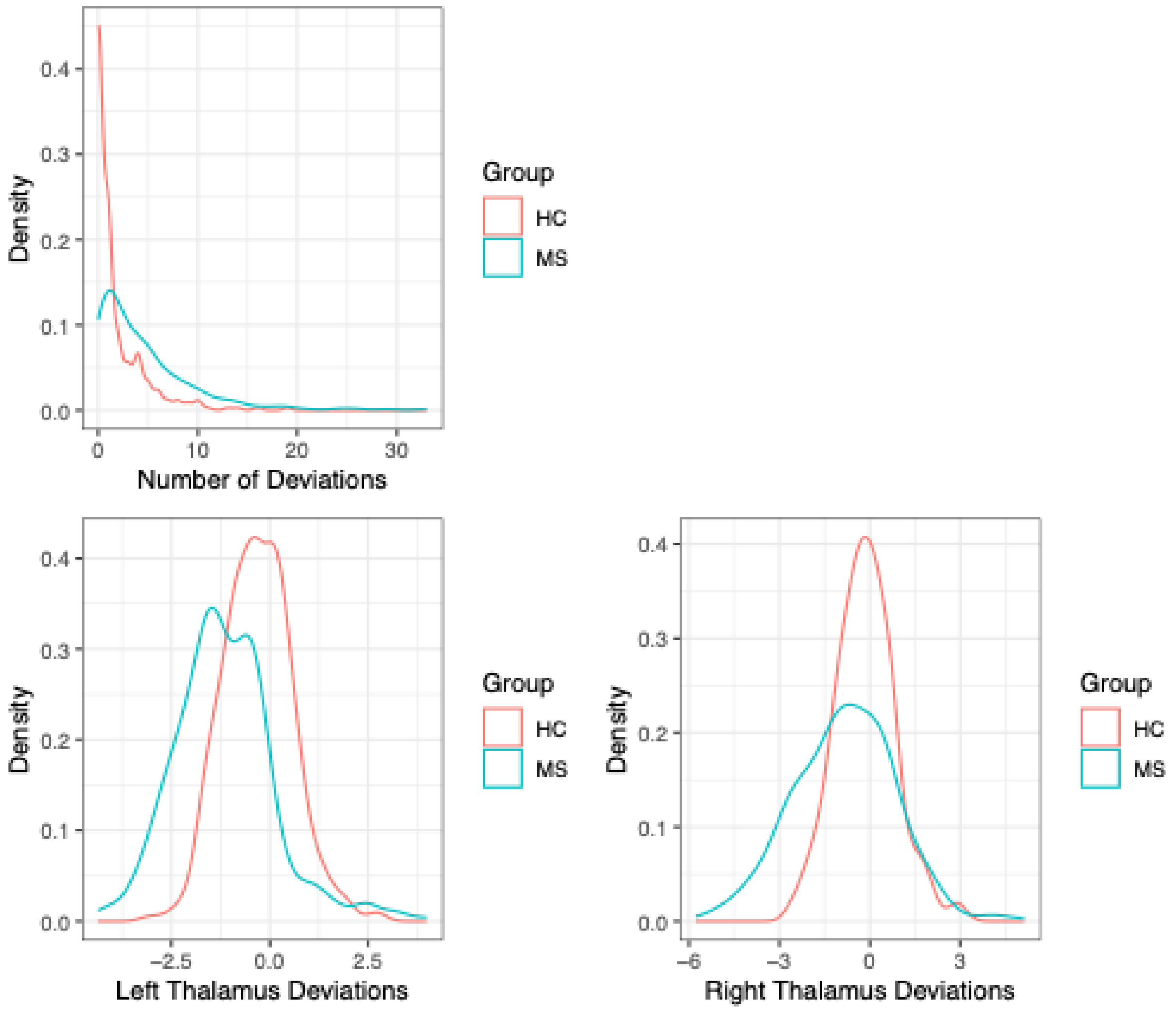


Top row: As reported in the main text, differences between the number of extreme deviations were significant between MS 4.51 (SE 4.95) HC 1.67 (SE 2.67), Cohen's d=0.71, 95% CI 0.56 to 0.87, p<0.0001. Bottom row: For the strongest effects, we show thalami Z-distributions. In pwMS, average deviations were $Z_{left}$=-1.16 and $Z_{right}$=-0.84, and for HC $Z_{left}$=-0.31 and $Z_{right}$=-0.15, being significantly different at Cohen's $d_{left}$ = 0.77, 95% CI 0.62 to 0.93], p < 0.00001) and Cohen's $d_{right}$ = 0.50, 95% CI 0.35 to 0.65], p < 0.00001).

***Supplemental Figure 2.*** **Region-averaged Z-values in MS and HC**

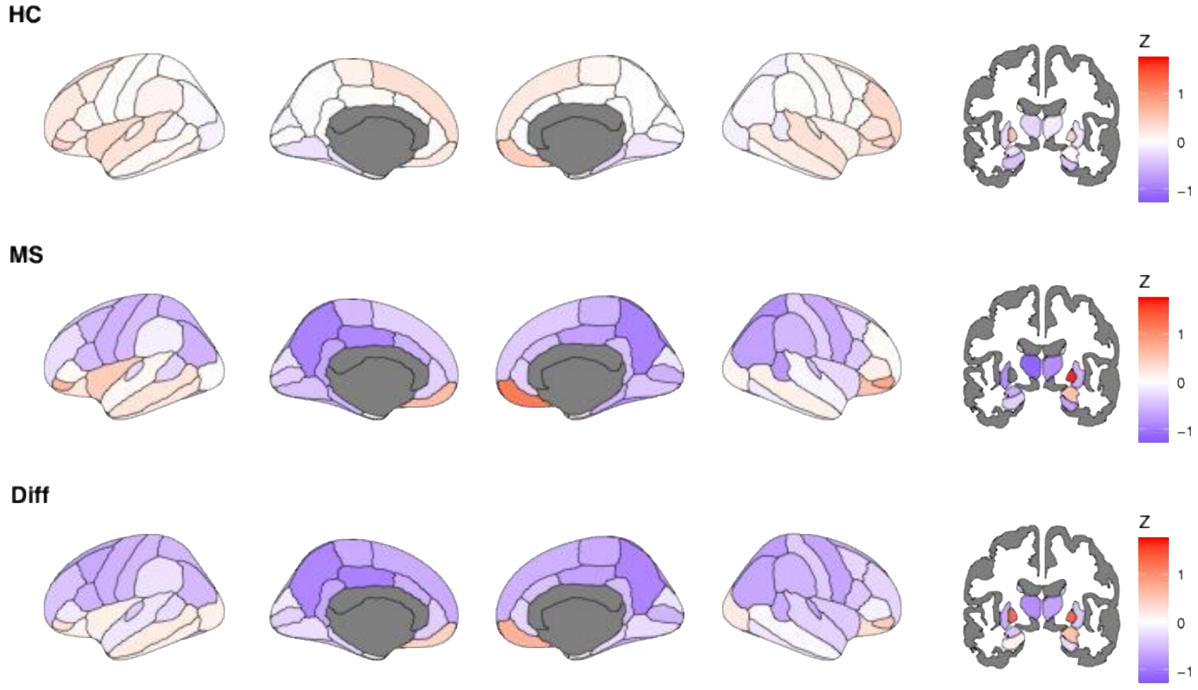


Regional Z-scores indicating deviations from the age-specific reference values in healthy controls (HC), multiple sclerosis (MS), and the delta between the examined samples. HC = healthy controls, MS = multiple sclerosis, Diff = difference between HC and MS.

Positive Z-scores in pallidum potentially originate from poor model performance as indicated by small variance in volumes explained by predictions during validation on matched healthy controls (see Supplemental Figure 7).

***Supplemental Figure 3.*** **Overlap of significant deviations indicating lower brain volumes (Z<-1.96) across participants at baseline for harmonised data**

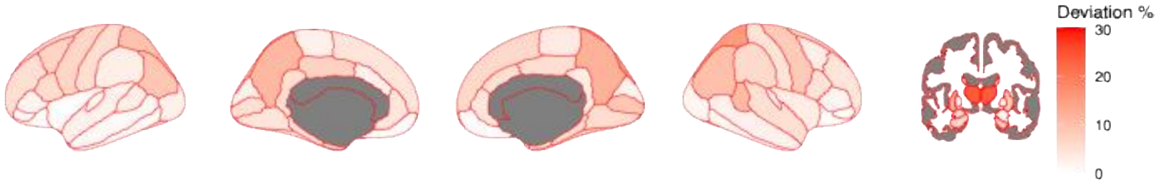

***Supplemental Figure 4.*** **Region-averaged Z-values in MS and HC for harmonised data**

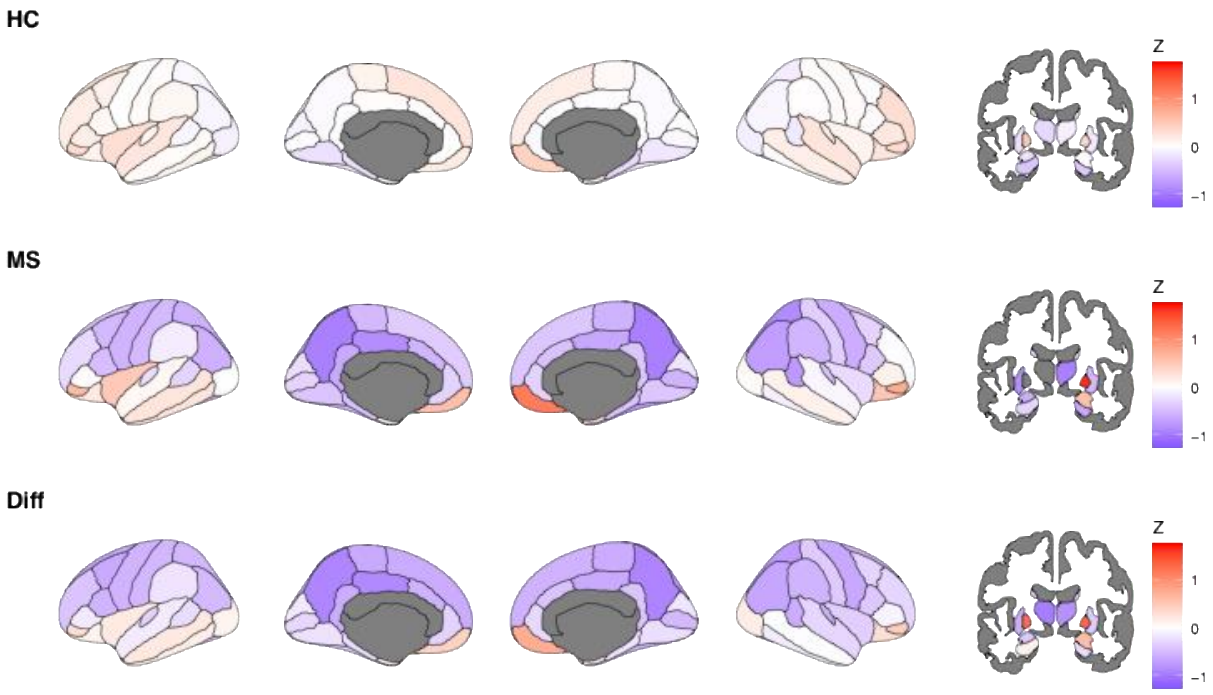


Regional Z-scores indicating deviations from the age-specific reference values in healthy controls (HC), multiple sclerosis (MS), and the delta between the examined samples. HC = healthy controls, MS = multiple sclerosis, Diff = difference between HC and MS.

Positive Z-scores in pallidum potentially originate from poor model performance as indicated by small variance in volumes explained by predictions during validation on matched healthy controls (see Supplemental Figure 7).

*Supplemental Figure 5:* **Overlap of significant deviations indicating lower brain volumes (Z<-1.96) across participants at baseline for harmonised data**

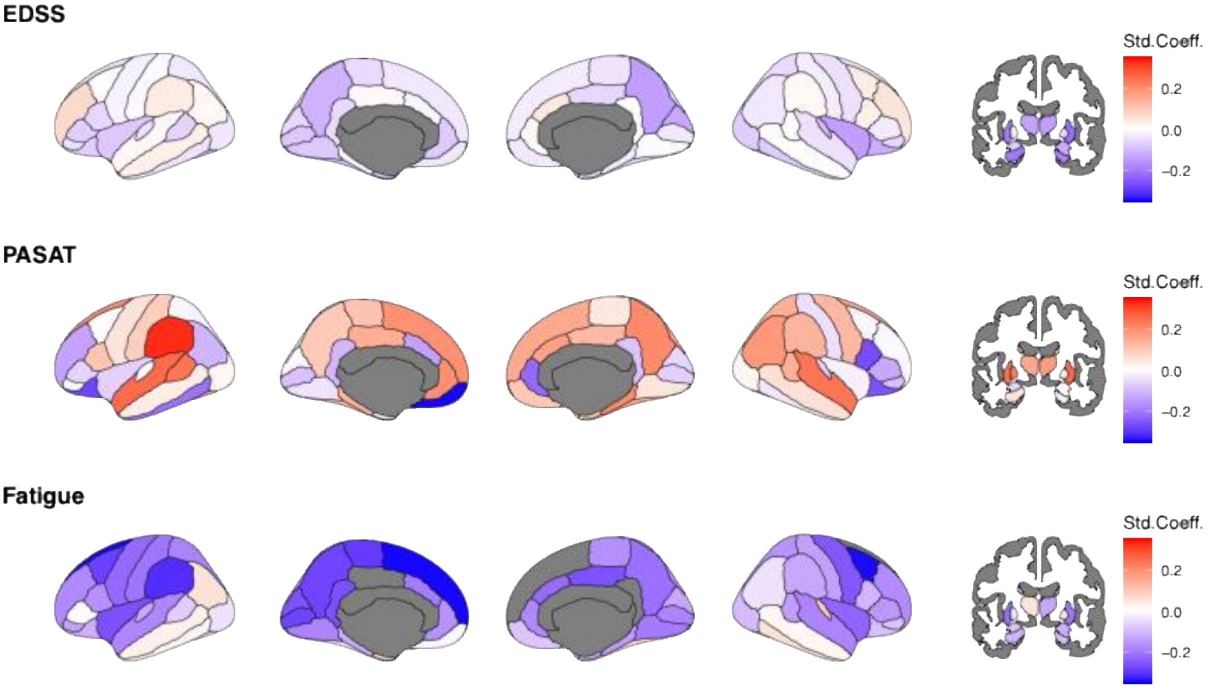


Std.Coeff. = standardized coefficient. Age and EDSS scores were available for N=214. Due to the harmonisation protocol leading to additional exclusions, tests were not possible for PASAT and FSS scores. The results generally correspond with the results observed in the main text, especially for associations of EDSS and subcortical volumes.

*Supplemental Figure 6.* **Longitudinal association of age, disability (EDSS), cognitive performance (PASAT), and fatigue on regional Z-scores for harmonised data**

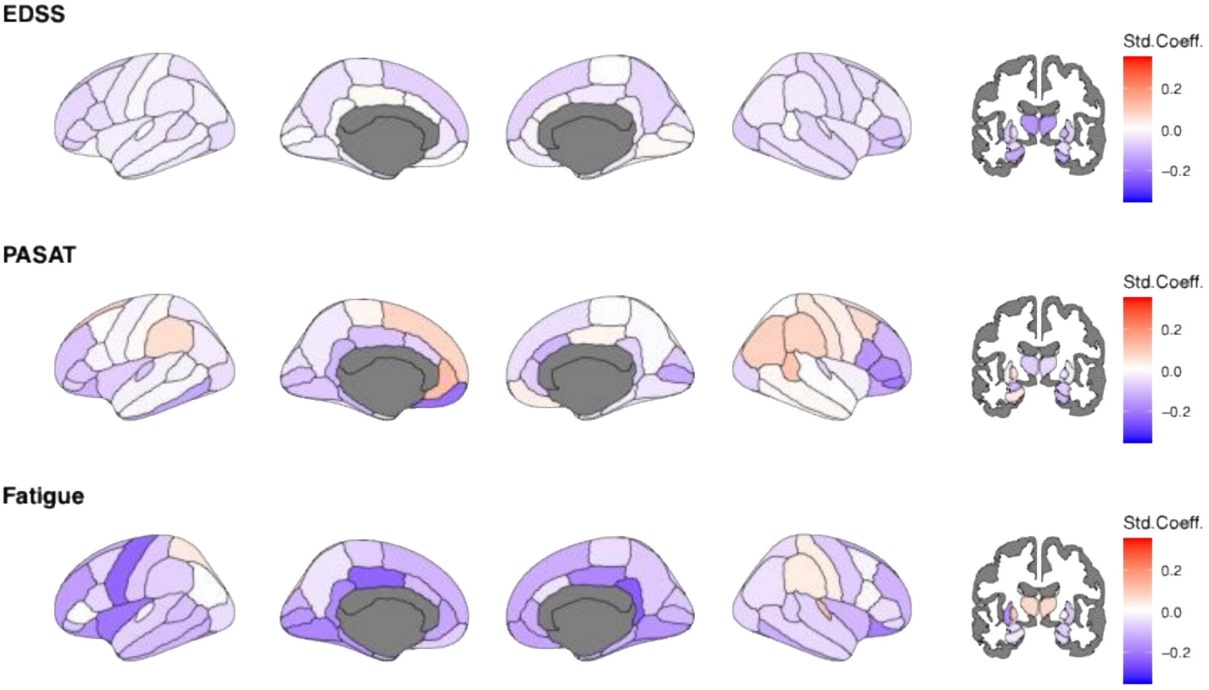


Std.Coeff. = standardized coefficient. Age and EDSS were available for 879 sessions (N=360). PASAT and fatigue scores were only available for N=95 (148 sessions) and N=133 (236 sessions), respectively. The high level of missingness did not allow imputations.

***Supplemental Figure 7.*** **Model performance: Variance explained and correlation coefficients for training (internal) and propensity matched (external) data.**

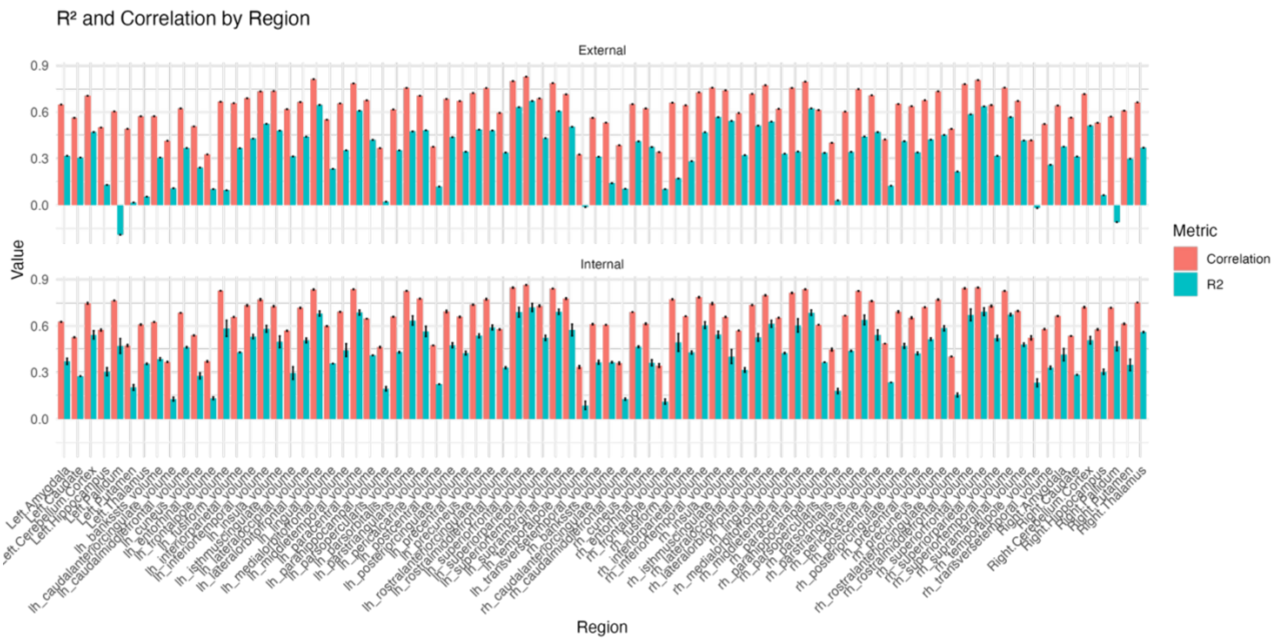


External data refer to the N = 351 propensity-matched healthy controls used for prediction. Internal data refers to the training data (N = 62 414).

***Supplemental Figure 8.*** **Model performance: Root mean squared error and mean absolute error for training (internal) and propensity matched (external) data.**

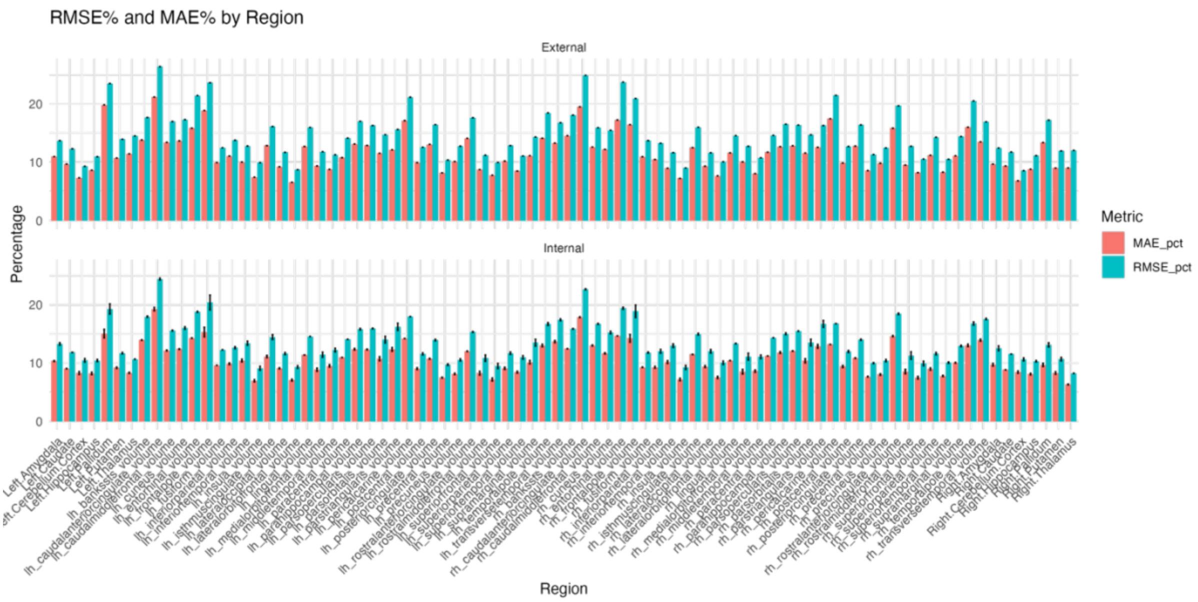


External data refer to the N = 351 propensity-matched healthy controls used for prediction. Internal data refers to the training data (N = 62 414).

*Supplemental Figure 9.* **Individual-level Z-scores for minimum and maximum EDSS and disease duration**

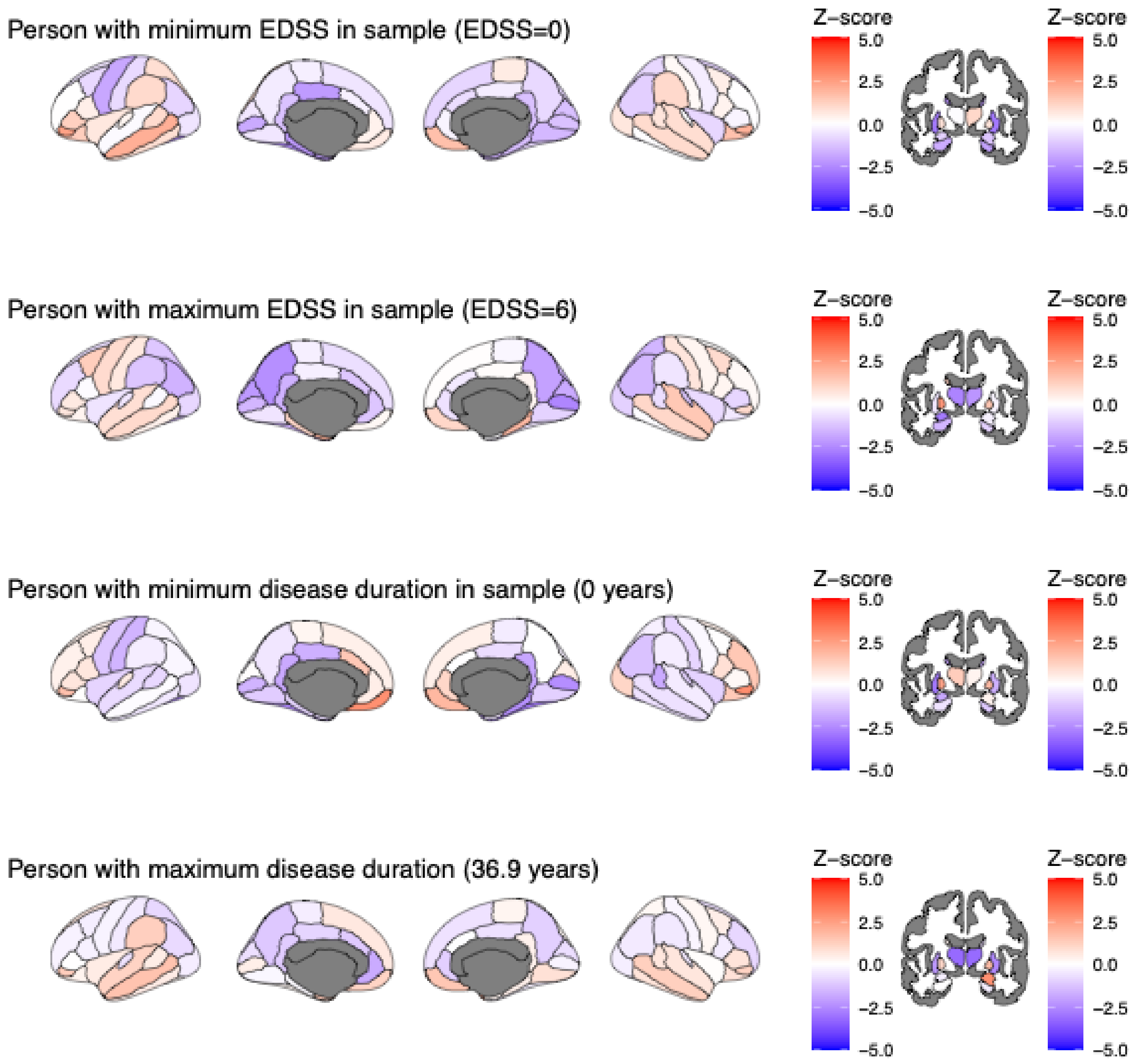

*Supplemental Figure 10.* **Individual-level Z-score-based extreme deviations for minimum and maximum observed EDSS and disease duration**

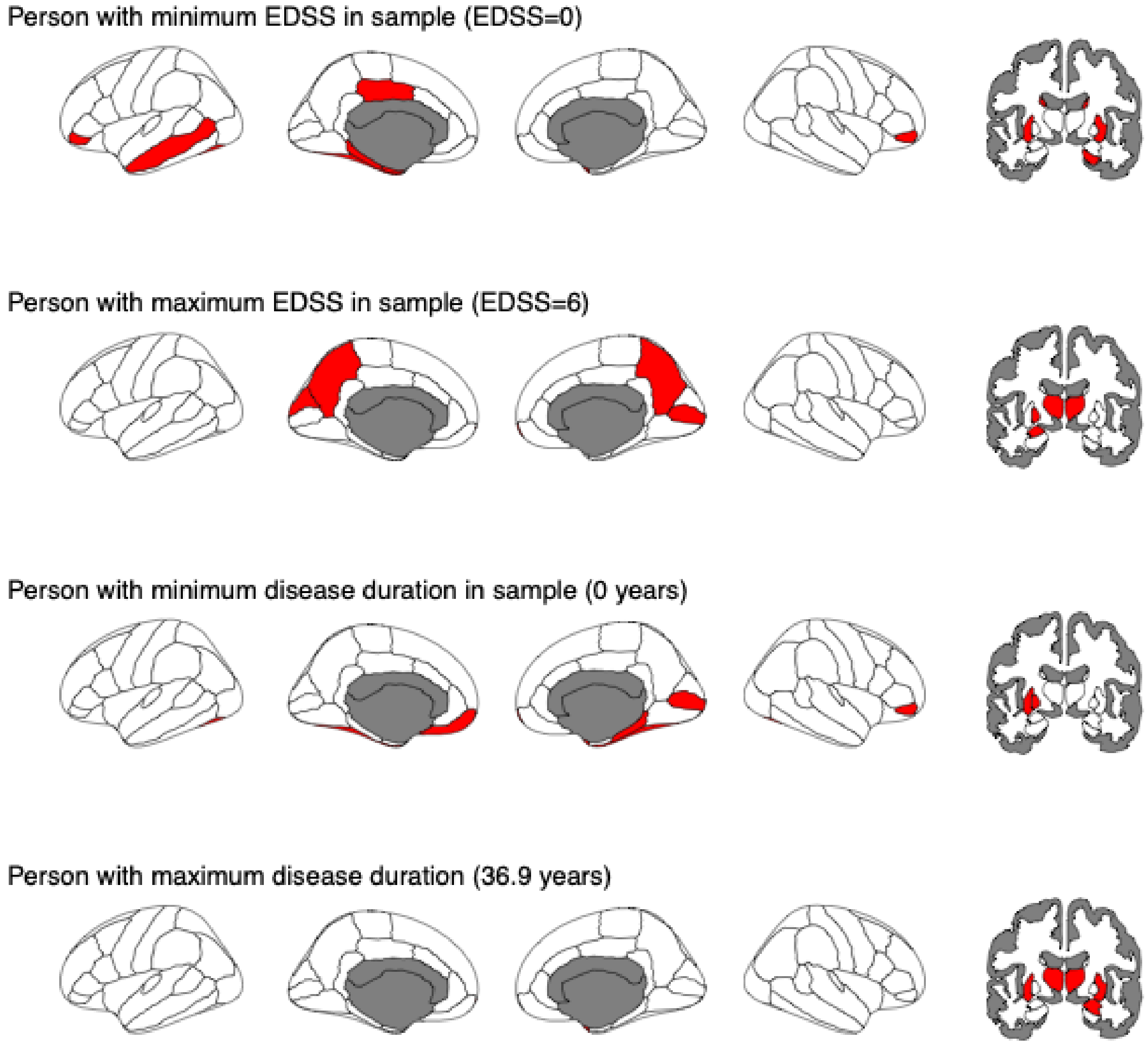


Extreme deviations were defined as $Z \leq -1.96$.

***Supplemental Figure 11.*** **Distribution of Z-scores at baseline in people with MS**

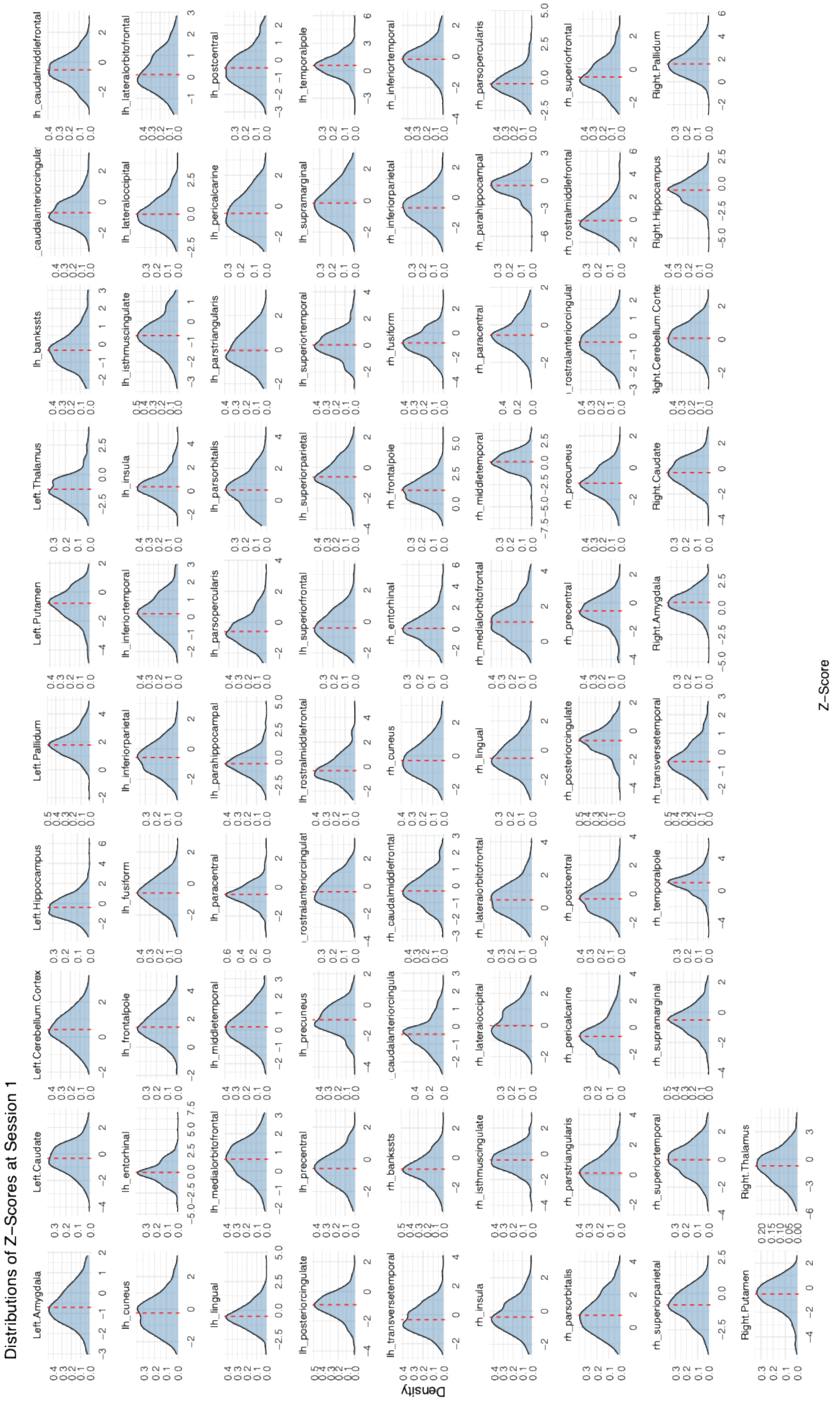

***Supplemental Figure 12.*** **Individual-level Z-scores for minimum and maximum EDSS and disease duration** ***using harmonised test data***

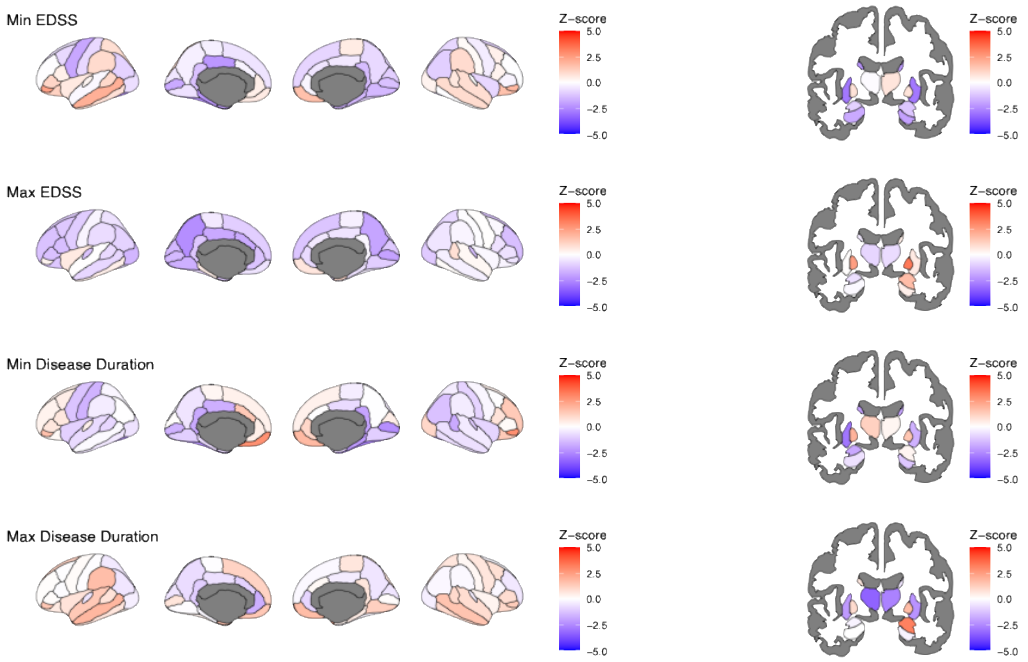

***Supplemental Figure 13.*** **Individual-level Z-score-based extreme deviations for minimum and maximum observed EDSS and disease duration *using harmonised test data***

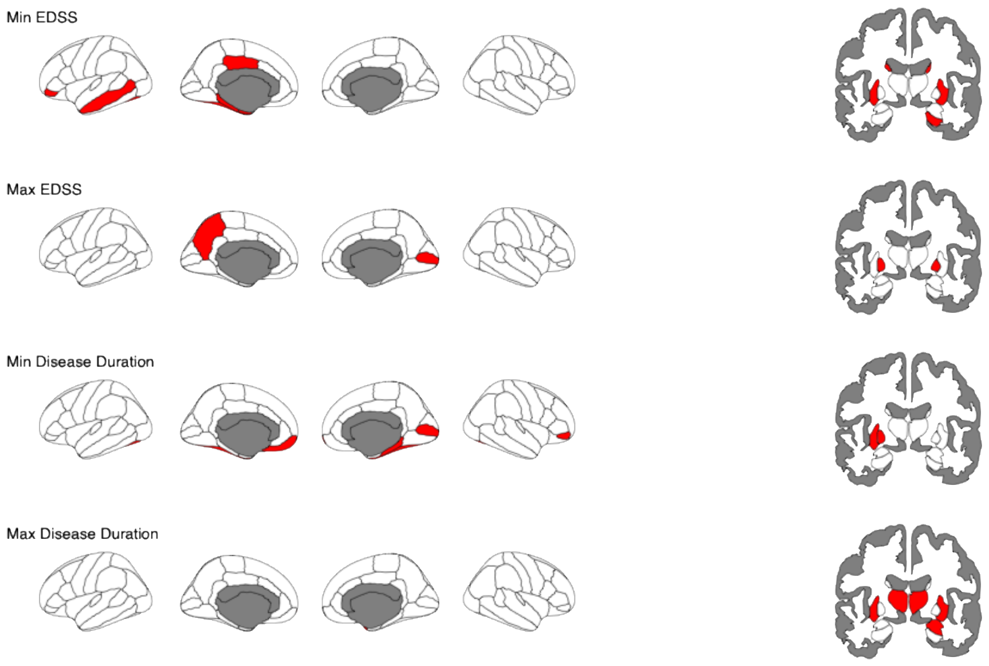

### Supplemental Notes

***Supplemental Note 1.*** **Ethics Approvals**

IRBs: **ABCD**: A centralised institutional review board approval of procedures was obtained from the University of California, San Diego. Written informed consent was obtained by a parent or guardian, and assent from the participants, before partaking in the ABCD study. **AddNeuroMed**: no IRB number available. **ADNI**: Data used in the preparation of this article were obtained from the Alzheimer’s Disease Neuroimaging Initiative (ADNI) database (adni.loni.usc.edu). The ADNI was launched in 2003 as a public-private partnership, led by Principal Investigator Michael W. Weiner, MD. The primary goal of ADNI has been to test whether serial magnetic resonance imaging (MRI), positron emission tomography (PET), other biological markers, and clinical and neuropsychological assessment can be combined to measure the progression of mild cognitive impairment (MCI) and early Alzheimer’s disease (AD). For up-to-date information, see www.adni-info.org. **HCP**: Ethics approval was obtained by the Norwegian Ethics Commission REK 567301, PVO 17/21624. Data used in the preparation of this work were obtained from the **Human Connectome Project (HCP)** database (https://ida.loni.usc.edu/login.jsp). The HCP project (Principal Investigators : Bruce Rosen, M.D., Ph.D., Martinos Center at Massachusetts General Hospital; Arthur W. Toga, Ph.D., University of Southern California, Van J. Weeden, MD, Martinos Center at Massachusetts General Hospital) is supported by the National Institute of Dental and Craniofacial Research (NIDCR), the National Institute of Mental Health (NIMH) and the National Institute of Neurological Disorders and Stroke (NINDS). HCP is the result of efforts of co-investigators from the University of Southern California, Martinos Center for Biomedical Imaging at Massachusetts General Hospital (MGH), Washington University, and the University of Minnesota. **Rockland Sample**: Ethics approval was obtained by the Norwegian Ethics Commission REK 567301, PVO 17/21624. **TOP**: The ethics approval was obtained by the Norwegian Ethics Commission REK 567301, PVO 17/21624. **UK Biobank**: Data were used under application 27412. Ethics approval was obtained from the National Health Service National Research Ethics Service (# 11/NW/0382).
An umbrella ethics approval (accross datasets) was obtained by the Norwegian Ethics Commission REK 567301, PVO 17/21624.

***Supplemental Note 2.*** **Longitudinal changes in training data with available longitudinal sessions.**

To provide an understanding of how Z-values change over time in healthy controls, we estimated Z-scores in training data with longitudinal data available. These data included N = 2,395 participants from the (UK Biobank = 2,163) and the Rockland sample (N = 232), with a mean±SD follow-up duration of 2.26±0.86 years.

Across this time, healthy controls had an increase in extreme deviations of 1.33±1.68 across the 82 examined regions (1.38±2.05%), which was a lower number of changes than the examined changes in the MS cohort (2.96±1.47%). These changes were defined as a transition from not-lower-than extremely negative reference values ($Z > -1.96$) towards extremely negative deviations ($Z \leq -1.96$), which indicates accelerated atrophy beyond what would be expected from the normative models.

***Supplemental Note 3.* Quality control analyses: associations between a) relapses, lesions, and b) Z-scores and extreme deviations.**

For quality control, we assessed the a) relationships between relapses and both the Z-scores as well as the total number of extreme deviations, as well as b) the relationships between both b1) T1-weighted as well as b2) T2-weighted lesions and both the Z-scores as well as the total number of extreme deviations. These associations were controlled for disease duration, using linear fixed effects regression models with Z-scores or the total number of extreme deviations being the predictors. Finally, we used the same parameters to explain EDSS scores and evaluated model performance at the inclusion or exclusion of key parameters using Likelihood Ratio Tests (LRT) and considered parameter estimates.

*Associations of norm deviation, relapses and lesions*

We did not observe a significant effect of the number of deviations on the number of relapses within 12 months prior baseline in the N = 118 participants with available data on the number of relapses ($p = 0.564$). We could neither observe an effect on the relapse rate over time ($p = 0.474$). None of the regional Z-scores were significantly associated with the number of relapses after FDR correction for multiple testing.

The number of T1-weighted lesions was significantly associated with the number of deviations (standardized coefficient = 0.28, 95% CI from 0.18 to 0.38, $p < 0.000001$) across participants (N = 364) indicating 0.09 added T1-weighted lesions with every added extreme deviation. Similarly, several regional Z-values were significantly associated with the number of T1-weighted lesions (see figure below).

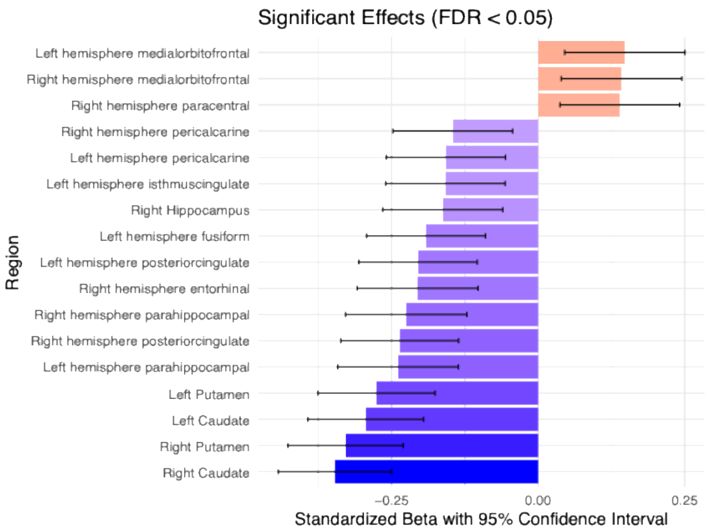


The number of T2-weighted lesions was however not associated with the number of extreme deviations at baseline ($p=0.516$). Yet, after FDA-correction region-level associations between T2-weigthted lesions and Z-scores in the right thalamus (standardized coefficient = -0.30, 95% CI from -0.47 to -0.12, $p_{FDR} = 0.0469$) and left superior frontal lobe (standardized coefficient = -0.31, 95% CI from -0.47 to -0.16, $p_{FDR} = 0.0095$) remained significant.

*Predictors of EDSS*

Finally, as a follow-up of the significant association between extreme deviations and T1-weighted lesions, we ran hierarchical regression models to examine the influence of either the number of T1-weighted lesions compared or the number of extreme deviations on EDSS. For that, we compared a full model predicting EDSS from T1-weighted lesions, the number of extreme deviations and the disease duration using LRT. First, T1-weighted lesions did not significantly predict EDSS, in any of the models (with ($p = 0.974$) or without ($p = 0.187$) including the number of extreme deviations in the model). Hence, removing T1-weighted lesions from the full model, did not significantly change model performance (LRT: $F = 0.001$, $p = 0.974$), and even indicated an increase in model performance, considering explained variance by $R^2 = 0.3\%$. However, removing the number of extreme deviations from the full model significantly lowered model performance ($F = 21.17$, $p = 0.000006$) from 9.09% of the variance explained to 3.95%. Standardized regression coefficients in the full model for the number of deviations were 0.24, 95% CI [0.14, 0.34], and for T1w lesions 0.0017, 95% CI [-0.10, 0.11]. Note that the variance inflation factor was below 1.1 across parameters across models, indicating the absence of multicollinearity.

For completeness, we also estimated models where EDSS was predicted by the number of extreme deviations, disease duration and either the number of T2-weighted lesions or the number of relapses at baseline. Both T2-weithted lesions ($p = 0.8696$ in the full model and $p = 0.9622$ in the model without number of extreme deviation) and the number of baseline relapses lesions ($p = 0.9557$ in the full model and $p = 0.944$ in the model without number of extreme deviation) were non-significant predictors of EDSS.

***Supplemental Note 4.*** **Differences between main and sensitivity analyses.**

While slight variability in results from analyses considering harmonised compared to non-harmonised test data can be expected, we report here the findings where statistical tests would indicate different conclusions in sensitivity analyses compared to the analyses reported in the main text.

*Magnitude of norm-deviations and clinical associations in cross-sectional analyses*. Sensitivity analyses indicated that only hippocampi (left: $\beta_{EDSS}$=–0.20; $p_{FDR}$=0.014; right: $\beta_{EDSS}$=–0.19; $p_{FDR}$=0.021) and putamen (left: $\beta_{EDSS}$=–0.20, $p_{FDR}$=0.014, right: $\beta_{EDSS}$=–0.22, $p_{FDR}$=0.009) were significantly associated with EDSS, not thalami ($p_{FDR}$>0.05), in the cross-sectional modelling, where EDSS was predicted from region-level Z-scores considering disease duration as a fixed covariate. No regional deviation was significantly associated with processing speed (PASAT score; $p_{FDR}$>0.05), but superior frontal volume deviations were significantly associated with fatigue (FSS; $\beta_{FSS}$ = -0.47; $p_{FDR}$ = 0.014), when using the same model structure but predicting PASAT or FSS instead.

***Supplemental Note 5.*** **Sensitivity analyses using Z < -2.33 as threshold to define critical deviations (CDs)**

*CD incidence*
The incidence ratio of CDs occuring was 3.47 95% CI [2.64, 4.58] comparing MS (2.38±3.43 CDs) to healthy control participants (0.69±1.57 deviations).

*Associations of critical deviations and clinical scores*
Using a more stringent threshold ($Z < -2.33$) to define CDs resulted in generally fewer deviations (see figure).

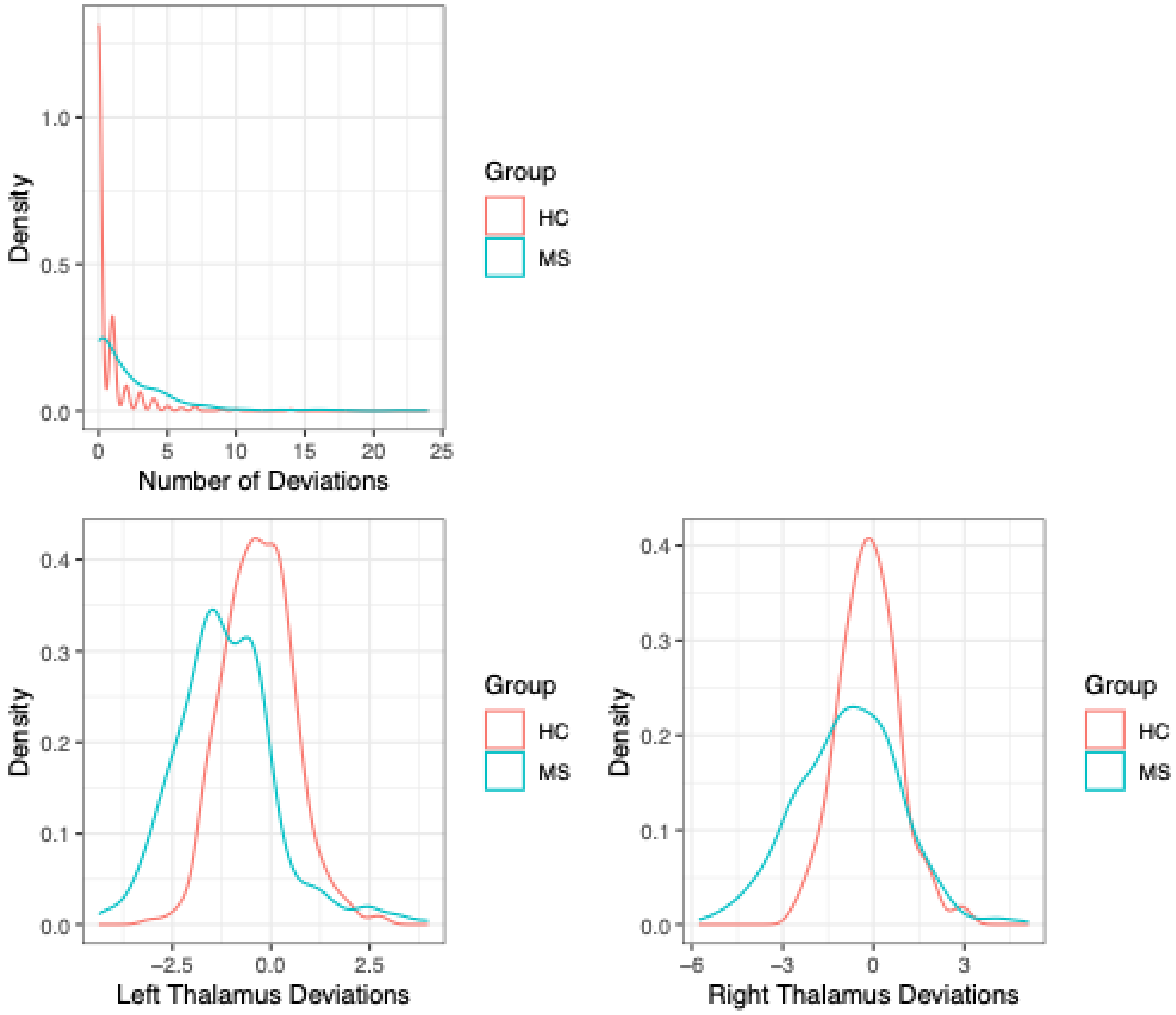


Moreover, the standardized longitudinal association between EDSS and the number of CDs $\beta_{EDSS} = 0.05$ 95% [-0.01, 0.10] assessed in a random intercept model controlling for disease duration was not significant $p = 0.078$, but remained nearly unchanged for the fixed effect cross-sectional linear model $\beta_{EDSS} = 0.24$ 95% CI [0.14, 0.34].

None of the associations with PASAT or FSS were significant.

Disease duration was only related to the CDs longitudinally $\beta_{DD} = 0.10$ 95% CI [0.04, 0.16].

*CD Profile*
Similar to the main analyses, thalami (20% and 17%) and the superior parietal cortex (10%) presented the largest number of people with MS having CDs in the same brain region. Instead of the pericalcarine, the putamen (8% and 7%) and fusiform area (8%) ranked third among the regions with the largest CD overlap across people (see figure).

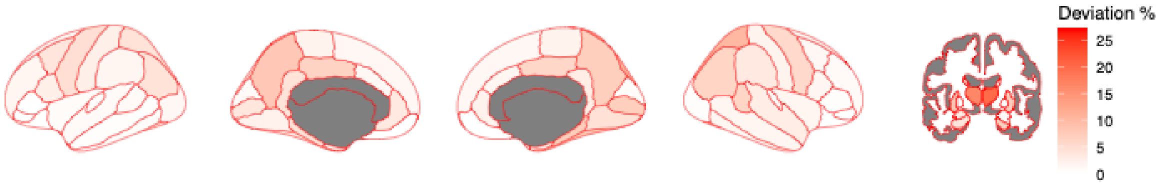


*Stratification*

All stratification results closely resemble the original analyses when using the same stratification regions (thalamus, superior parietal cortex, pericalcarine, see figure), but risk individuals in the risk group had higher EDSS over time in a random intercept model ($\beta_{EDSS}$ = 0.29 95% CI [0.15; 0.42]) than in the original analyses (repetition from main text: $\beta_{EDSS}$ = 0.13 95% CI [0.03; 0.24])

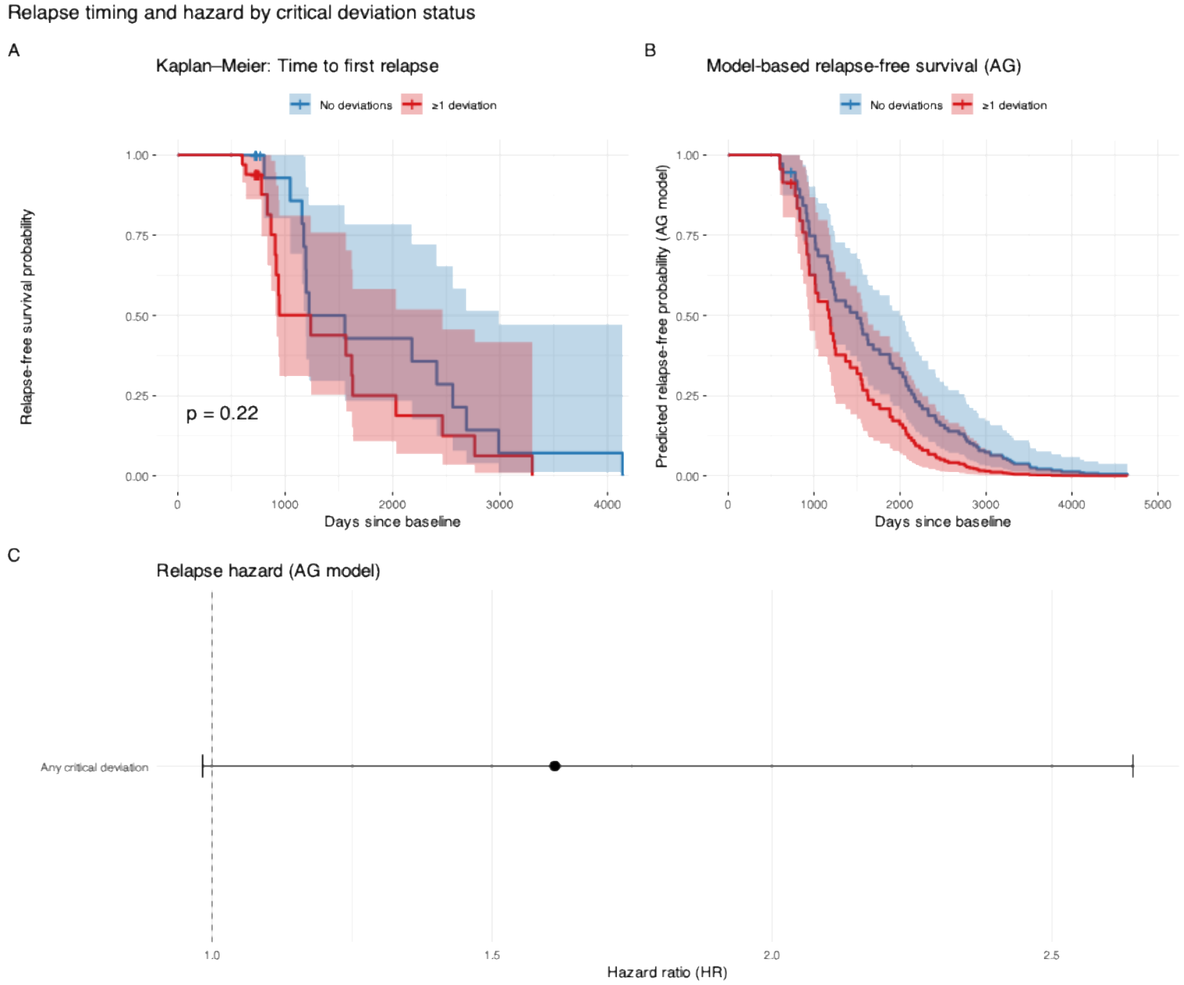

***Supplemental Note 6.* Scanners and protocols in the training cohort**

A total of 52 unique scanners was used in the acquisition of the training data. For an overview of each sub-cohort see Supplemental Table 1.

The ABCD[2] study used a total of 27 scanners from Siemens, GE, and Philips at 3T all using a harmonised protocol that can be found here: https://abcdstudy.org/images/Protocol_Imaging_Sequences.pdf

AddNeuroMed data was acquired from six different 1.5T MR systems (four General Electric, one Siemens and one Picker). Acquisition protocols were compatible with the Alzheimer disease neuroimaging initiative (ADNI) protocol.[9]

ADNI data was available from 13 scanners (2 Phillips, 3 GE, 8 Siemens) at 3T. Acquisition protocols can be found here: https://adni.loni.usc.edu/data-samples/adni-data/neuroimaging/mri/

Human Connectome Project[5] (HCP)[6] acquisition was done using a Siemens Trio, Verio, or Skyra 3T using TR=2400ms, TE=2.14, FA=8°, with detailed protocols available at https://www.humanconnectome.org/storage/app/media/documentation/s1200/HCP_S1200_Release_Appendix_I.pdf

Rockland Sample[7] acquisition was done on a single Siemens Trio 3T, and the protocol can be found here: https://rocklandsample.org/for-researchers-rockland-sample-ii-neuroimaging-protocol

Thematically Organized Psychosis (TOP) data were acquired at one Siemens 1.5T Sonata: TR=2730ms, TE=3.93ms, FA=7°, GE 3T Signa HDxT: TR=7.8ms, TE=2.956ms, FA=12° (one subset with HNS coil, one subset with 8HRBRAIN coil), and a GE 3T Discovery GE750: TR=8.16ms, TE=3.18ms, FA=12°.

UK Biobank[8] data were acquired using the same scanner model, Siemens 3T Skyra, with TR=2000ms, TE=2.01ms, FA=8° at 3 sites. The detailed protocol is available here: https://www.fmrib.ox.ac.uk/ukbiobank/protocol/
